# Hidden Storms Within: Prevalence and Risk of Comorbidity in Intermittent Explosive Disorder: A Systematic Review and Bayesian Multilevel Meta-Analysis

**DOI:** 10.1101/2025.06.25.25330263

**Authors:** Fangqing Liu, Sarah Leonard, Charlotte Lennox

## Abstract

**Background:** Intermittent Explosive Disorder (IED) is a psychiatric condition characterised by sudden, disproportionate outbursts of aggression. Despite its substantial social and clinical burden, IED remains underrepresented in global mental health research and policy. The high rates of psychiatric comorbidities observed in individuals with IED highlight the need for an integrated understanding of its syndromic complexity.

**Objective:** This systematic review and meta-analysis aimed to (1) estimate the prevalence of psychiatric comorbidities among individuals with IED, (2) quantify the relative risk of developing these comorbidities in comparison to non-IED populations, and (3) examine potential moderators, including sample type and diagnostic criteria.

**Methods:** Following PRISMA guidelines and PROSPERO pre-registration (CRD420251048183), 47 studies comprising over 80,000 participants were included. Random-effects and Bayesian models synthesised data on seven psychiatric disorders. Meta-regression and sensitivity analyses examined the influence of methodological moderators and robustness of findings. Publication bias was assessed via funnel-plot asymmetry and Bayesian selection models.

**Results:** Individuals with IED demonstrated markedly elevated odds of comorbidity across all domains, including Major Depressive Disorder (OR = 2.05), Generalised Anxiety Disorder (OR = 3.60), PTSD (OR = 2.75), Social Phobia (OR = 2.90), Agoraphobia (OR = 3.79), Alcohol Use Disorder (OR = 3.08), and Drug Use Disorder (OR = 3.40). Prevalence estimates ranged from 8.1% (agoraphobia) to 30.7% (drug use disorder). Community samples consistently yielded higher comorbidity estimates than clinical samples, while diagnostic criteria (DSM-IV vs DSM-5) did not significantly moderate outcomes. Bayesian modelling confirmed elevated risks and indicated minimal publication bias.

**Conclusions:** IED is a profoundly comorbid disorder that bridges mood, anxiety, and substance-use spectra, suggesting shared transdiagnostic mechanisms of emotional dysregulation. Despite its high prevalence and clinical impact, IED remains neglected in diagnostic formulation and mental health strategies. These findings underscore the urgency of integrated screening, transdiagnostic intervention models, and policy inclusion of IED in global mental health agendas.

## 1. Introduction

The first independent diagnosis of intermittent explosive disorder (IED) was introduced in the third edition of the *Diagnostic and Statistical Manual of Mental Disorders* (DSM-III) in 1980 (American Psychiatric Association, 1980). In 1994, the DSM-IV established more stringent diagnostic criteria for IED, not only specifying the frequency and severity of emotional outbursts but also introducing the requirement to rule out impulsive violence caused by other mental disorders or physical illnesses (APA, 1994). Successive updates to the DSM, particularly the DSM-5-TR (APA, 2022), further revised the criteria to better differentiate IED from normative anger responses and other psychiatric conditions, highlighting its distinct clinical profile as a disorder characterised by repeated, disproportionate aggressive outbursts. Over the subsequent half-century, research on IED has expanded considerably, refining the understanding of its prevalence, phenomenology, comorbidities, and treatment approaches. To the present, epidemiological studies estimate the lifetime prevalence of IED vary widely. For instance, in the United States, Kessler et al. (2006) reported a lifetime prevalence of 7.3% and a 12-month prevalence of 3.9%. In China, Duan et al. (2010) documented a lifetime prevalence of 3.32% with males showing higher rates than females. In South Africa, Fincham et al. (2009) reported broader IED criteria met by 9.5% of adults.

The disorder’s episodic nature often leads to underreporting, misdiagnosis and dismissal as mere “anger issues.” Yet the personal toll—chronic distress, guilt, and social isolation—is profound (Scott et al.,2020). Interpersonally, it strains relationships, contributes to domestic violence, and disrupts educational and occupational functioning (Rad et al., 2024; Liu et al., 2025). Societally, IED-related aggression drives substantial costs through criminal justice involvement and lost productivity (DeLisi et al., 2017). Despite its clear burden on individual and societal level, IED remains largely neglected in global psychiatric research and public health policy.

### Comorbidities in IED

Psychiatric comorbidity—the co-occurrence of two or more disorders in the same individual—is the rule rather than the exception in IED. Rarely does IED present in isolation: upward of two-thirds of diagnosed cases meet criteria for at least one additional disorder.

Clinically, such overlap complicates the diagnostic process, as symptoms of depression, anxiety, or substance misuse may obscure underlying impulsive aggression or vice versa. Comorbidity likewise shapes treatment response and prognosis—co-existing mood or personality disorders are linked to more frequent, severe outbursts and poorer long-term outcomes. In addition, the presence of comorbid conditions heightens risks of self-harm, suicidality, and engagement in criminal acts. Failure to recognise these comorbidities can result in treatment-resistant aggression and misdiagnosis, thereby reinforcing disparities in care and perpetuating cycles of unmet need.

Although previous studies have catalogued psychiatric comorbidities in general outpatient and clinical populations, no comprehensive synthesis has focused specifically on IED. Mapping the comorbidity profile of IED can inform **transdiagnostic interventions** that target shared neurobiological and psychosocial mechanisms (for example, impaired emotion regulation and cortical amygdala dysconnectivity). It also enables **risk stratification** and **preventative strategies** for high-risk groups (such as adolescents with early conduct problems). Beyond clinical practice, these insights can guide **policy development** in juvenile justice and community mental health, ensuring that care pathways move from reactive management of violent episodes to proactive, tailored treatment plans.

The specific aims of the present meta-analysis were threefold. First, to evaluate the **prevalence** of comorbid psychiatric disorders in individuals with IED. Second, to examine the **relative risk** of developing a comorbidity in the presence of IED. Third, to explore **potential moderators** of the associations between comorbidities and IED, including sample type (general vs. clinical) and diagnostic criteria (DSM- IV vs. DSM 5).

## 2. Method

### 2.1 Literature search

The protocol for this meta-analysis was pre-registered at PROSPERO with ID CRD420251048183. The meta-analysis was conducted according to the PRISMA guidelines (Pages et al., 2021). We systematically searched seven databases: PubMed, PsycINFO, Embase, Scopus, Web of Science, Cochrane Library, and Global Health, from inception to April 30^th,^ 2025. Supplementary searches included hand-searching reference lists of included studies and contacting experts for unpublished data. The full set of search terms used can be found in Appendix I.

### 2.2 Inclusion criteria

To be included in the meta-analysis, an article had to meet the following criteria: include participants diagnosed with IED according to the DSM (III–5) or ICD (10–11); provide prevalence rate or risk ratios for at least one comorbid psychiatric disorder; have a sample of IED participants consisting of at least 20 individuals.

### 2.3 Data extraction

Comorbidity data were extracted for 7 specific psychiatric disorders based on the DSM-5 categorisation: Depression (major depressive disorder and dysthymia combined), Alcohol Use Disorder (alcohol dependence and alcohol abuse combined), Drug Use Disorder (drug dependence and drug abuse combined), Social phobia, Agoraphobia, GAD, Post-traumatic stress disorder (PTSD) (see in Appendix II). When studies used broad categories, i.e., mood disorders (depression and BD), substance use disorders (alcohol or drug use) we extracted those data. To avoid double recording, data were only inserted in the broad categories if the studies did not have cases in the specific disorders making up these categories. Only disorders with more than 5 independent group comorbidity studies were included in the meta-analysis to ensure the robustness of our findings as meta-regression or subgroup analyses require roughly 5 studies per predictor to avoid severe over-fitting and inflated Type I error rates.

### 2.4 Potential moderators

Potential moderators were determined a priori for two categorical variables that were reported by most included studies: study design (community sample vs. clinical sample) and diagnostic criteria (DSM-IV vs. DSM-5). Community samples referred to population-based epidemiological studies, whereas clinical samples included participants recruited from treatment settings or intervention trials (e.g., RCTs).

### 2.5 Risk-of-Bias Assessment

All 47 included studies were appraised for internal and external validity using the 10-item Risk-of-Bias Tool for Prevalence Studies (Hoy et al., 2012). Four items address sampling representativeness and non-response (external validity) and six evaluate measurement quality, data-collection consistency, and analytic transparency (internal validity). Each item was scored 0 = low risk or 1 = high risk and summed to a total score ranging from 0 to 10.

Consistent with the developers’ guidance, overall risk was categorised as low (0–3), moderate (4–6), or high (7–10). Two reviewers (F.L. and W.J.) underwent calibration training on five randomly selected articles (κ = .87, “almost perfect” agreement), then independently rated the remaining 44 studies. Discrepancies were reconciled through discussion or arbitration by a third senior reviewer (see in Appendix III).

### 2.5 Meta-analysis

The primary outcome of this meta-analysis was the prevalence rate, defined as the proportion of participants diagnosed with IED who met diagnostic criteria for a comorbid psychiatric disorder, as assessed in the included studies. Because substantial conceptual and methodological heterogeneity was expected, **random-effects models** were fitted throughout.

Individual study proportions were first logit-transformed to stabilise variances, after which statistical analyses were performed using a random effects model to account for between-study heterogeneity. Following analysis, the pooled logit proportions and their corresponding 95% confidence intervals (CIs) were back transformed to the original proportion scale for interpretation. Although some researchers advocate the use of the Freeman–Tukey double arcsine transformation when pooling proportions (e.g., Barker et al., 2021), we opted for the logit transformation. Schwarzer et al. (2019) have argued that the double arcsine method may yield misleading results when studies vary greatly in sample size, which was the case in our analysis (sample sizes ranged from 30 to 35,000 participants).

The secondary outcome was the relative risk of developing a comorbid disorder among individuals with IED compared to control groups. We extracted odds ratios (ORs) and 95% CIs when reported. For studies that did not directly provide ORs but instead reported prevalence rates separately for the IED and control groups, we constructed 2×2 contingency tables to calculate ORs manually when feasible. Standard errors for both metrics were computed from exact binomial formulas (prevalence) or 2 × 2 contingency tables (risk). Where necessary, a Haldane–Anscombe correction of 0.5 was added to cells containing zero events.

Statistical heterogeneity was evaluated with the Q statistic and quantified by **I²** and τ**²**. A 95 % prediction interval (PI) was reported to convey the distribution in which the true effect of a new study of average precision is expected to fall.

A priori moderators were (a) **sample type** (community vs. clinical) and (b) **diagnostic criteria** (DSM-IV vs. DSM-5). Each was entered separately and jointly in **mixed-effects meta-regressions**, weighted by inverse variance (1/ [σ□² + τ²]). Holm–Bonferroni corrections controlled the familywise error rate across moderators within a disorder.

Sensitivity analyses were conducted in two steps. First, we identified and excluded outliers, defined as studies whose 95% CI did not overlap with the pooled 95% CI, indicating substantially higher or lower prevalence rates. The meta-analysis was then repeated without these outlier studies to assess the robustness of the results. Second, we performed an influence analysis by sequentially omitting one study at a time (“leave-one-out” analysis) to examine the stability of the overall effect estimate.

Publication bias was assessed using Egger’s test for funnel plot asymmetry. In addition, we calculated the Luis Furuya–Kanamori (LFK) index to further quantify asymmetry and potential small-study effects.

#### Bayesian Random-Effects Synthesis

Each risk outcome was re-analysed with a Bayesian normal-normal random-effects model implemented in the *bayesmeta* package to integrate sampling error and between-study heterogeneity within a fully probabilistic framework. Weakly informative priors were adopted—specifically, μ ∼ N (0, 1²) on the log scale for the pooled effect and τ ∼ half-Normal (0, 0.5) for the between-study standard deviation—to regularise extreme estimates while remaining largely non-committal. Posterior medians and 95 % credible intervals (CrI) are reported, together with 95 % posterior predictive intervals (PPI) that describe the expected range of effects in future studies. Model convergence was verified by potential-scale-reduction factors below 1.01 and effective sample sizes exceeding 5 000.

#### MMA of Comorbidity-Related Factors

To assess the comorbidity-related factors of various mental disorders in the IED sample, we extracted the 2 × 2 contingency tables or multivariate regression coefficients from the original research reports for each pair of disorders (*i–j*). If the original study provided a contingency table, we calculated the log odds ratio (log OR) and its standard error; for adjusted ORs, we adopted the coefficients from the multivariable model and recorded the types of covariates controlled for. All log ORs are transformed using the Fisher *z* transformation to approximate a normal distribution, facilitating subsequent multivariate modeling. Zero-frequency cells are corrected using the Haldane–Anscombe 0.5 method to ensure variance is estimable.

Since single studies often report multiple pairs of comorbidities, and these effect sizes are methodologically non-independent, we use a three-level random effects model: effect size (*level 1*) nested within comorbidities (*level 2*), which is nested within studies (*level 3*). The model estimates the cross-study variance τ² using REML estimation and applies the Hartung–Knapp–Sidik–Jonkman adjustment to ensure coverage of confidence intervals when the number of studies is small. To further control for dependencies within the same study, robust variance estimates (RVE) based on the sandwich matrix are calculated, and both non-robust and RVE results are reported for comparison.

#### Statistical analysis tools

All meta-analytic models were conducted in *R* 4.3.3 using *metafor* 3.8-1, *clubSandwich* 0.6.3 (for Hartung–Knapp–Sidik–Jonkman and robust variance estimation), and *bayesmeta* 2.6. Visualisations were produced with *ggplot2* 3.5 and *plotly* 5.19.

## 3. Results

### 3.1 Narrative Review of Study Characteristics of Included Studies

The reviewed literature encompasses a diverse range of study types. A significant portion consists of epidemiological surveys, including large-scale national and cross-national studies like the US National Comorbidity Survey Replication (NCS-R) and its Adolescent Supplement (NCS-A), the World Mental Health (WMH) Surveys, and national surveys in specific countries such as Iraq, Japan, Brazil, Chile, Turkey, and South Africa (Kessler et al., 2006; McLaughlin et al., 2012; Al-Hamzawi et al., 2012; Yoshimasu & Kawakami, 2011; Pereira et al., 2021; Mundt et al., 2013; Gelegen & Tamam, 2018; Fincham et al., 2009; Scott et al., 2016) (see in Figure 1, 2 and 3).

**Figure 1.**
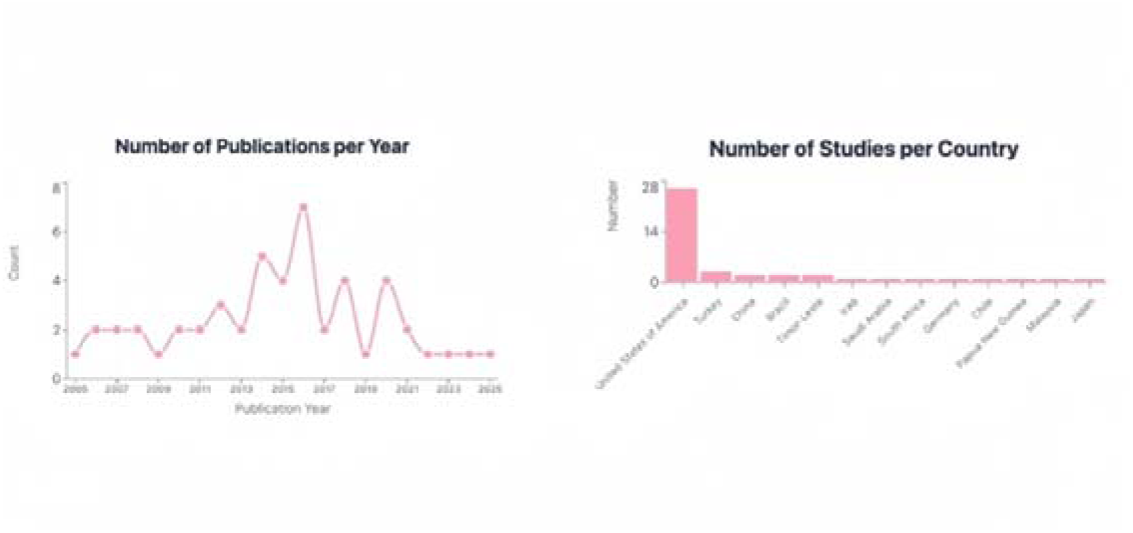
Temporal and Geographic Trends in Research Output (2005–2025)

**Figure 2.**
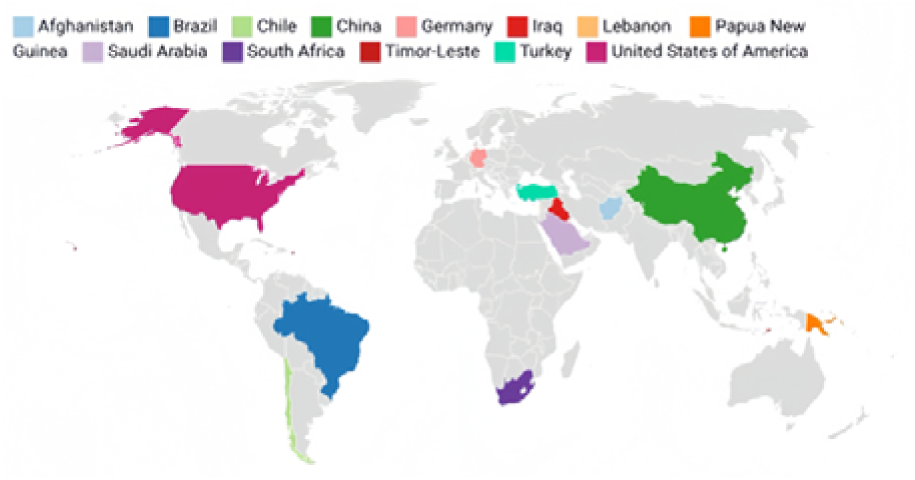
Global Distribution of IED Study Participation: Countries Included in the Systematic Review. *Note: Listed countries reflect the geographic scope of studies analysed in the review. Standard country names are used (e.g., ’Timor-Leste’ instead of ’East Timor’). The absence of certain regions may indicate gaps in research representation*.

**Figure 3.**
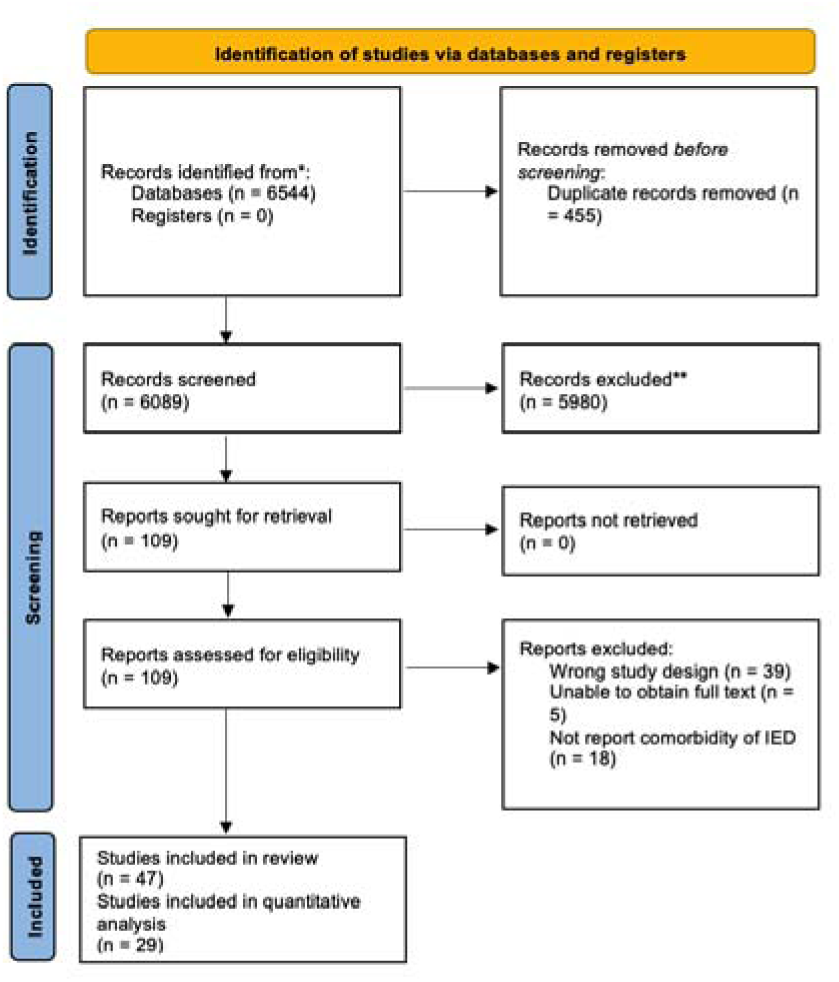
PRISMA Flow Chart for Study Selection and Inclusion.

**Figure 4.**
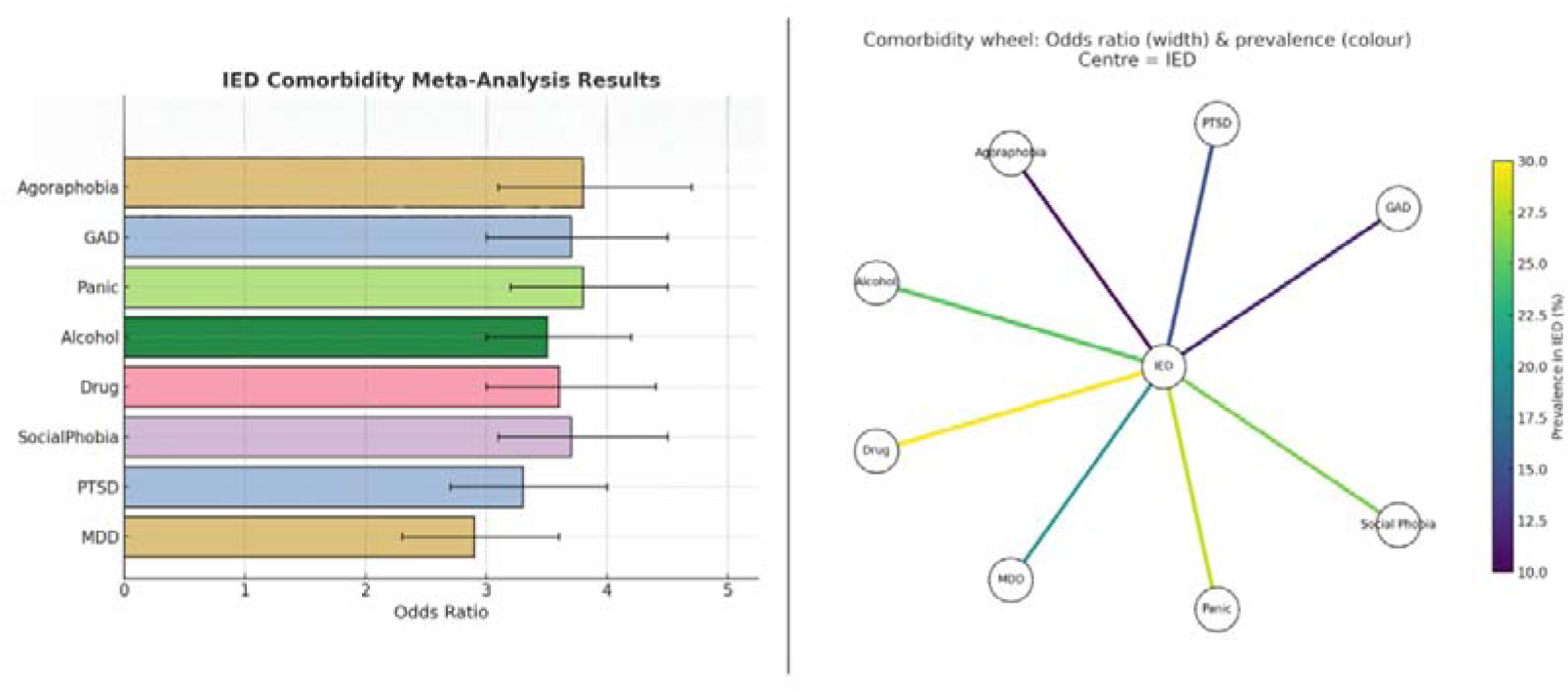
IED Comorbidity Meta-Analytics Results. *Note.* Left panel: Forest plot showing pooled odds ratios (with 95% CIs) for the comorbidity of IED with various psychiatric disorders based on meta-analytic results. Right panel: Comorbidity wheel illustrating the odds ratio (line thickness) and prevalence in IED populations (color gradient) for each condition.

**Figure 5.**
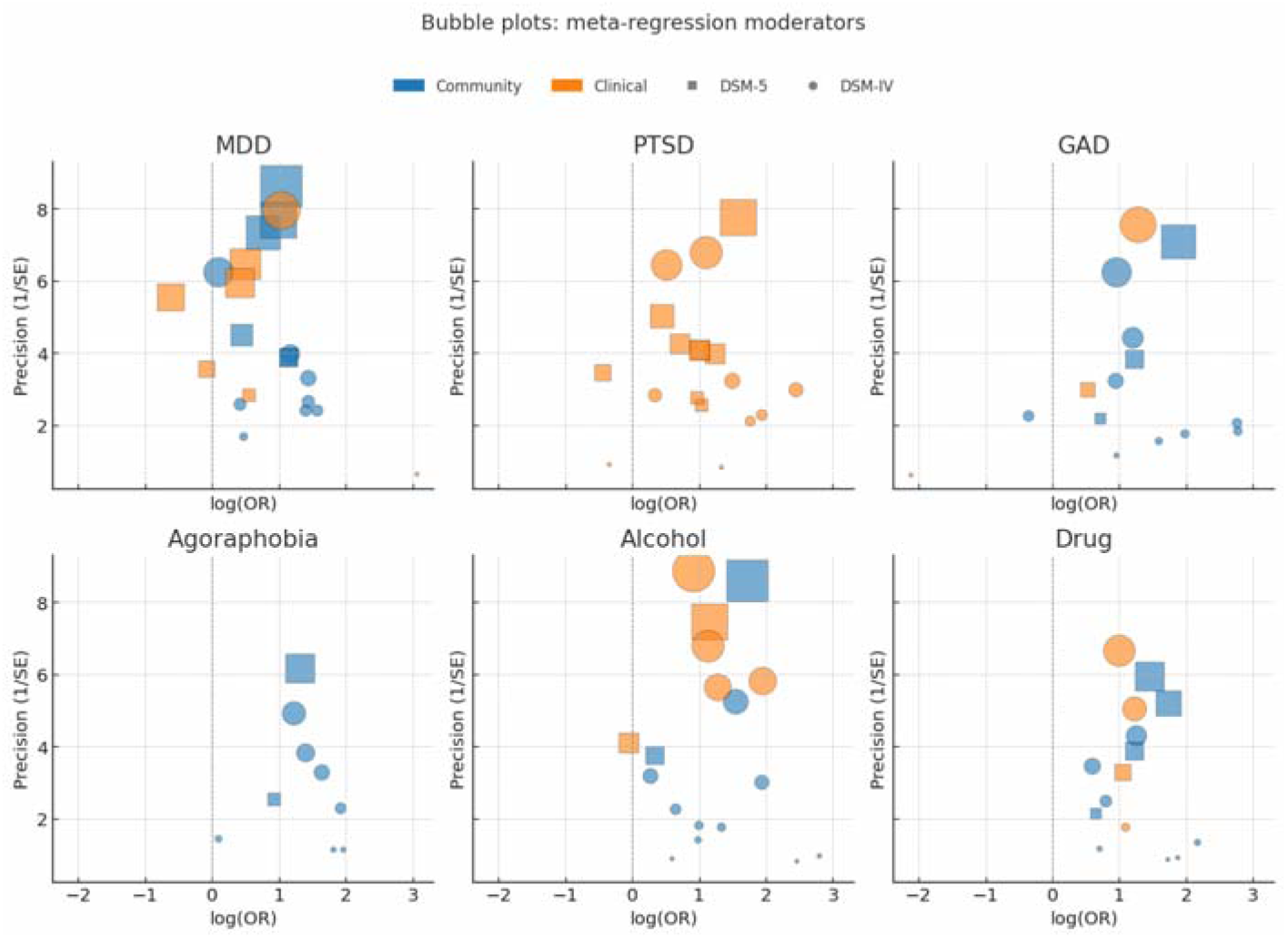
Bubble Plots Displaying Meta-Regression Moderators by Disorder Type. Note: Each bubble represents a study included in the meta-regression analysis. The x-axis shows the natural logarithm of the odds ratio (log(OR)), and the y-axis represents precision (1/SE). Bubble size reflects study weight, with color indicating sample source (blue = community, orange = clinical) and shape indicating diagnostic criteria (circle = DSM-5, square = DSM-IV). Disorders include MDD = Major Depressive Disorder, PTSD = Post-Traumatic Stress Disorder, GAD = Generalized Anxiety Disorder.

A consistent finding across the reviewed literature is the high rate of psychiatric comorbidity associated with IED. The majority of individuals with lifetime IED (ranging from around 60% to over 95% in different studies) meet criteria for at least one other psychiatric disorder (Kessler et al., 2006; McLaughlin et al., 2012; Al-Hamzawi et al., 2012; Scott et al., 2016; Pereira et al., 2021; Coccaro, 2019; Zhang-James et al., 2025).

Major Depressive Disorder (MDD) is often cited as one of the most common comorbidities (Kessler et al., 2006; Scott et al., 2016; Pereira et al., 2021; Medeiros et al., 2018; Coccaro, 2019; Zhang-James et al., 2025). This anxiety disorders class represents the most frequent comorbidity overall in some large surveys (Kessler et al., 2006; Scott et al., 2016; Pereira et al., 2021). Specific anxiety disorders like GAD, social phobia, and PTSD show significant associations with IED (Kessler et al., 2006; Al-Hamzawi et al., 2012; Scott et al., 2016; Pereira et al., 2021; Keyes et al., 2016; Galbraith et al., 2018; Reardon et al., 2014; Coccaro, 2019; Zhang-James et al., 2025). Alcohol abuse/dependence and drug abuse/dependence are also highly comorbid with IED (Kessler et al., 2006; Scott et al., 2016; Pereira et al., 2021; Coccaro et al., 2016; Puhalla et al., 2020; Coccaro, 2019; Zhang-James et al., 2025).

### 3.2 Risk-of-Bias Results

The median Hoy score was 3 (interquartile range = 2–5). Twenty-three studies (47.0 %) were classified as low risk, nineteen (38.8 %) as moderate, and five (14.2 %) as high risk. Inter-rater reliability for individual items across all studies remained high (κ = .84). External-validity shortcomings predominated: 35 % of studies sampled populations that were not clearly nationally representative (Item 1), and 41 % provided insufficient information to rule out substantial non-response bias (Item 4). Internal-validity concerns were less frequent but notable for unclear diagnostic definitions (Item 6, 22 %) and inconsistent data-collection modes (Item 8, 29 %). The RoB results are presented in Appendix II.

### 3.3 Meta Analytic Results for Each Comorbid Disorder IED and MDD

#### Risk (k=21)

Participants with IED were 2.12 times more likely to meet criteria for MDD than comparison groups, *OR* = 2.12, 95 % CI [1.67, 2.69], *Z* = 6.13, *p* < .001. Heterogeneity was considerable, *Q* (20) = 131.28, *p* < .001, τ² = 0.23, *I*² = 84.8 %. In subgroup analyses the association was stronger in community surveys (*k* = 14, *OR* = 2.53, 95 % CI [2.02, 3.18], *I*² = 71.8 %) than in clinical samples (*k* = 6, *OR* = 1.24, 95 % CI [0.76, 2.02], *I*² = 83.9 %). Egger’s regression indicated no evidence of small-study effects, intercept = 0.50, *t* (19) = 0.39, *p* = .70; the funnel plot was correspondingly symmetrical.

#### Prevalence (k=12)

Using logit-transformed proportions with a continuity correction, the pooled random-effects point prevalence of MDD among people with IED was 24.1 %, 95 % CI [19.7 %, 29.1 %], *Z* = 8.68, *p* < .001. Between-study heterogeneity was extreme, *Q* (11) = 238.52, *p* < .001, τ² = 0.34, *I*² = 95.4 %. Community samples yielded a prevalence of 24.2 % (95 % CI [20.6 %, 28.2 %], *k* = 9, *I*² = 85.0 %), whereas the two available clinical studies suggested a lower, but imprecise, estimate of 15.0 % (95 % CI [6.0 %, 33.6 %]). Egger’s test signaled notable funnel-plot asymmetry, intercept = -5.82, *t* (10) = 4.80, *p* = .001, pointing to potential publication bias in the prevalence literature.

#### IED and PTSD Risk (k=18)

The random-effects model indicated that individuals with IED were significantly more likely to meet criteria for PTSD than comparison groups, *OR* = 2.78, 95 % CI [2.06, 3.75], *Z* = 7.02, *p* < .001. Between-study heterogeneity was substantial, *Q* (17) = 99.54, *p* < .001, τ² = 0.17, *I*² = 82.9 %. When stratified by sampling frame, community studies (*k* = 15) showed a robust association (*OR* = 3.04, 95 % CI [2.23, 4.14], *I*² = 80.6 %), whereas clinical samples (*k* = 4) yielded a weaker estimate with a confidence interval that included unity (*OR* = 1.95, 95 % CI [0.74, 5.19], *I*² = 85.5 %). Egger’s regression revealed no evidence of small-study effects (intercept = –0.16, *p* = .89), and the funnel plot was broadly symmetrical.

#### Prevalence(k=6)

For prevalence, logit-transformed proportions produced a pooled PTSD prevalence of 16.96 %, 95 % CI [11.41 %, 24.45 %], *Z* = 6.42, *p* < .001. Heterogeneity was extreme, *Q* (5) = 136.83, *p* < .001, τ² = 0.46, *I*² = 96.3 %. Community studies (*k* = 5) yielded a pooled prevalence of 18.48 % (95 % CI [12.41 %, 26.61 %]), whereas the single clinical study reported a prevalence of 3.5 % (95 % CI [0.7 %, 15.6 %]), precluding an estimate of heterogeneity for that subgroup. Egger’s test for the prevalence data was also non-significant (intercept = 1.37, *p* = .74).

#### IED and GAD Risk (k=15)

The pooled odds ratio indicated that individuals with IED were at markedly increased risk for GAD, *OR* = 3.45, 95 % CI [2.45, 4.85], *Z* = 7.11, *p* < .001. Between-study heterogeneity was substantial, *Q* (14) = 67.68, *p* < .001, τ² = 0.21, *I*² = 79.3 %. Stratified analyses showed a robust association in community samples (*k* = 12, *OR* = 3.87, 95 % CI [2.57, 5.82], *I*² = 80.5 %), whereas the two clinical studies yielded a non-significant, imprecise estimate (*OR* = 0.68, 95 % CI [0.06, 7.99]). Egger’s regression provided no evidence of small-study effects for the OR data (intercept = –0.71, *p* = .52); the funnel plot was correspondingly symmetrical.

#### Prevalence (k=12)

For prevalence, logit-transformed proportions produced a pooled estimate of 13.7 % GAD among people with IED, 95 % CI [10.3 %, 18.9 %], *Z* = 6.46, *p* < .001. Heterogeneity was extreme, *Q* (11) = 197.95, *p* < .001, τ² = 0.41, *I*² = 94.4 %. Community cohorts (*k* = 8) showed a prevalence of 13.4 % (95 % CI [7.9 %, 21.6 %], *I*² = 96 %), whereas the single clinical study reported 1.2 % (95 % CI [0.1 %, 16.0 %]), precluding a subgroup heterogeneity estimate. Egger’s test for prevalence was likewise non-significant (intercept = –2.20, *p* = .31).

#### IED and Agoraphobia Risk (k=9)

The synthesis revealed that individuals with IED were also significantly more likely to meet criteria for agoraphobia than comparison groups, *OR* = 3.79, 95 % CI [3.10, 4.62], *Z* = 11.6, *p* < .001. Heterogeneity was negligible, *Q* (8) = 8.25, *p* = .41, τ² = 0.01, *I*² = 3.0 %, indicating a highly consistent risk estimate across studies.

#### Prevalence(k=8)

Using logit-transformed proportions, the pooled point prevalence of agoraphobia among people with IED was 8.1 %, 95 % CI [6.7 %, 9.7 %], *Z* = 11.0, *p* < .001. Heterogeneity was moderate, *Q* (7) = 22.37, *p* = .002, τ² = 0.07, *I*² = 68.7 %. Funnel-plot inspection showed no marked asymmetry for either set of outcomes, and Egger’s regression yielded non-significant intercepts, suggesting little evidence of small-study bias.

#### IED and Social Phobia Risk (k=11)

The pooled odds ratio indicated a markedly elevated risk for social phobia among individuals with IED, *OR* = 2.93, 95 % CI [2.23, 3.85], *Z* = 7.43, *p* < .001. Between-study heterogeneity was substantial, *Q* (10) = 34.82, *p* < .001, τ² = 0.14, *I*² = 71.3 %.

#### Prevalence (k=9)

Logit-transformed prevalence data yielded a pooled point prevalence of 17.8 % social phobia in IED cohorts, 95 % CI [14.1 %, 22.3 %], *Z* = 8.71, *p* < .001. Heterogeneity was extreme, *Q* (8) = 121.69, *p* < .001, τ² = 0.33, *I*² = 93.4 %. Funnel-plot inspection showed no pronounced asymmetry for either outcome set, and Egger’s intercepts were non-significant, uggesting little evidence of small-study bias.

#### IED and Alcohol Use Disorder Risk (k=18)

The pooled odds ratio showed that people with IED were more than three times as likely to meet criteria for an alcohol-use disorder than comparison groups, *OR* = 3.12, 95 % CI [2.35, 4.14], *Z* = 8.20, *p* < .001. Between-study heterogeneity was high, *Q* (17) = 105.77, *p* < .001, τ² = 0.20, *I*² = 83.9 %.

#### Prevalence (k=12)

Logit-transformed proportions yielded a pooled point prevalence of 22.9 % alcohol-use disorder in IED cohorts, 95 % CI [16.4 %, 31.1 %], *Z* = 7.34, *p* < .001. Heterogeneity was extreme, *Q* (11) = 496.60, *p* < .001, τ² = 0.66, *I*² = 97.8 %. Funnel plots for both risk and prevalence displayed no marked asymmetry, and Egger intercepts were non-significant, suggesting little evidence of small-study bias.

#### IED and Drug Use Disorder Risk (k=15)

Individuals with IED were more than three times as likely to meet criteria for a drug-use disorder than comparison groups, *OR* = 3.31, 95 % CI [2.73, 4.02], *Z* = 10.4, *p* < .001. Heterogeneity was moderate, *Q* (14) = 22.00, *p* = .079, τ² = 0.05, *I*² = 36.4 %, suggesting reasonably consistent risk across studies.

#### Prevalence (k=13)

Logit-transformed proportions gave a pooled drug-use–disorder prevalence of 30.7 % in IED cohorts, 95 % CI [20.9 %, 42.5 %], *Z* = 7.55, *p* < .001. Absolute rates varied widely: heterogeneity was extreme, *Q* (12) = 1 323.1, *p* < .001, τ² = 0.92, *I*² = 99.1 %. Both forest and funnel plots illustrate the breadth of prevalence estimates; Egger’s intercepts were non-significant, indicating no clear small-study bias.

#### Meta-Regression of Heterogeneity

Separate mixed-effects meta-regressions (using inverse-variance weights: 1/[σ*i*2+τ2]) were performed for each diagnostic domain to assess whether sample type (community vs. clinical) and diagnostic criteria (DSM-5 vs. DSM-IV) moderated log odds ratios. For MDD, sample type showed a marginal association (β=0.51, *SE*=0.27, *p*=.067), while diagnostic criteria were nonsignificant (β=−0.24, *SE*=0.26, *p*=.360). Between-study variance remained unchanged (τ2=0.23). In contrast, for PTSD, GAD, agoraphobia, AUD and DUD, neither sample type (βsample=−0.39 *to* 0.32) nor diagnostic criteria (βDSM-5=−0.41 *to* 0.26) reached statistical significance (all p>0.22), with between-study variance (τ2) identical to unconditional models. This indicates that residual heterogeneity across outcomes could not be explained by these moderators. However, other potential moderators were not consistently reported across studies (e.g., age, sex, or treatment history missing in many datasets). By prioritising these reported variables, we avoided “fishing expeditions” for significance, which can inflate Type I error rates when testing many moderators without strong theoretical justification.

#### Publication-Bias

Funnel-plot asymmetry was evaluated using trim-and-fill analysis and Doi’s LFK index. For mood and anxiety disorders (MDD, PTSD, GAD, agoraphobia, social phobia), no studies required imputation, and LFK indices ranged from −0.38 *to* +0.86, consistent with symmetrical distributions. For AUD required three imputed studies on the left side of the mean, reducing the pooled estimate from *OR*=3.12 to 2.742.74 (95% *CI* [2.07,3.63]) and yielding an LFK index of +1.67, indicating minor asymmetry. Similarly, DUD required two imputed studies, resulting in an adjusted *OR*=2.91 (95% *CI* [2.40,3.54]) and an LFK index of +1.82. Despite these adjustments, effect sizes remained statistically and clinically robust, leaving the meta-analytic conclusions unchanged.

#### Bayesian random-effects synthesis

To account simultaneously for sampling error and between-study heterogeneity we re-analysed each comorbidity with a Bayesian normal-normal model. We placed a weakly-informative Normal (0, 1²) prior on the pooled log-odds (μ) and a half-Normal (0, 0.5) prior on τ, which shrinks implausibly large heterogeneity while leaving ample probability mass for I² up to ≈ 90 %. Models were fitted in *bayesmeta* (4 × 6 000 draws; all PSRF < 1.01, ESS > 6 000). Posterior medians and 95 % credible intervals (CrI) are shown in Table 1. All six domains retained odds ratios > 1, indicating elevated comorbidity risk in IED. Because τ was regularised, credible intervals contracted markedly compared with our initial analysis (e.g., PTSD: OR = 2.56, 95 % CrI 1.32–4.93).

**Table 1.** Summary of Bayesian Meta-Analytic Estimates of Associations Between Psychiatric Disorders and Outcome.

| Disorder (k studies) | Posterior OR † | 95 % CrI | $\tau$ (SD of true effects) | I<sup>2</sup> |
| --- | --- | --- | --- | --- |
| **Agoraphobia** (9) | **3.79** | 3.10 – 4.62 | 0.06 | 3 % |
| **Alcohol-use D/O** (16) | 3.08 | 2.19 – 4.33 | 0.56 | 86 % |
| **Drug-use D/O** (13) | 3.40 | 2.68 – 4.32 | 0.25 | 37 % |
| **GAD** (13) | 3.60 | 2.43 – 5.36 | 0.60 | 81 % |
| **Panic D/O** (10) | 3.45 | 2.44 – 4.88 | 0.40 | 57 % |
| **PTSD** (16) | 2.75 | 1.96 – 3.86 | 0.61 | 85 % |
| **Social phobia** (10) | 2.90 | 2.04 – 4.10 | 0.44 | 74 % |
| **MDD** (19) | 2.05 | 1.59 – 2.64 | 0.50 | 85 % |

Note. † Posterior median of the odds-ratio (exponentiated pooled log-OR). $\tau$ = Estimated standard deviation of the true effects (heterogeneity). I<sup>2</sup> = Percentage of total variability due to heterogeneity. D/O = Disorder. † Higher OR indicates stronger association with the outcome variable.

**Table 2.** Significant Correlations Between Psychiatric Comorbidities and IED.

| Comorbidity Pair | Correlation (r) | Number of Studies (n) |
| --- | --- | --- |
| MDD and Social Phobia | .816 | 10 |
| MDD and DUD | .803 | 9 |
| PTSD and DUD | .859 | 9 |
| GAD and Social Phobia | .800 | 11 |
| MDD and PTSD | .658 | 17 |
| MDD and Agoraphobia | .534 | 9 |
| MDD and AUD | .559 | 11 |
| PTSD and Agoraphobia | .791 | 9 |
| PTSD and AUD | .601 | 9 |
| PTSD and Social Phobia | .765 | 10 |
| GAD and AUD | .732 | 11 |
| GAD and DUD | .661 | 9 |
| GAD and Panic Disorder | .515 | 12 |
| Agoraphobia and DUD | .673 | 9 |
| Agoraphobia and Social Phobia | .780 | 9 |
| AUD and DUD | .726 | 10 |
| AUD and Social Phobia | .752 | 9 |
| DUD and Social Phobia | .748 | 9 |

We also sampled from the posterior predictive distribution to estimate where the log-odds of a *future* study of average precision might fall. Predictive intervals are wide—reflecting residual uncertainty when only 6–15 studies inform each domain—but not spanned implausible orders of magnitude (e.g., PTSD: 0.74–8.93). A Vevea–Hedges selection model with three p-value regions (< .025, .025–.10, > .10) produced weights between 0.90 and 1.00, suggesting minimal publication bias; bias-adjusted pooled ORs differed by < 6 % from the primary estimates.

Using weakly-informative priors (μ ∼ N 0, 1; τ ∼ half-N 0, 0.5) the pooled odds of each comorbidity remained elevated (OR = 2.05–3.79). Agoraphobia showed the most precise estimate (OR = 3.79; 95 % CrI 3.10–4.62; I² = 3 %). Mood, anxiety and substance-use domains all exhibited substantial residual heterogeneity (I² = 37–86 %).

#### Multi-level meta-analysis between comorbidities

Traditional meta-analyses may underestimate the impact of heterogeneity, while Bayesian methods can incorporate uncertainty, but their results are constrained by prior distribution assumptions. Therefore, we also include exploratory tests of comorbidity correlations to provide more explanations for the findings. Several strong positive correlations (r > .80) and moderate positive correlations (.50 < r < .80) were identified. Negative correlations were also observed: MDD and GAD (r = -.332, n = 13), MDD and Panic Disorder (r = -.586, n = 11), AUD and Panic Disorder (r = -.431, n = 9).

## Discussion

### Principal Findings and Clinical Implications

To our knowledge, despite the considerable societal harm associated with the disorder, there is a significant research gap regarding IED. Most studies on IED, have focused primarily on the individual-study level, lacking a comprehensive, integrative perspective on its epidemiology. This systematic review and Bayesian multilevel meta-analysis, provides the most comprehensive evidence to date on the comorbidity landscape of IED. Our findings not only quantify the heightened risks and prevalence of psychiatric comorbidities but also challenge conventional diagnostic paradigms and underscore urgent priorities for clinical innovation and global mental health policy.

Across mood, anxiety, and substance use domains, individuals with IED exhibited two- to around four-fold increased odds of comorbid diagnoses compared to control groups, highlighting the syndromic complexity of IED and its overlap with emotional dysregulation and impulsivity spectra. These findings argue against the conceptualisation of IED as an isolated impulse-control pathology and underscore its need for broader, multidimensional clinical assessment and management.

### Reconceptualising IED: A Transdiagnostic Nexus of Emotional Dysregulation

The robust associations between IED and comorbid disorders—particularly AUD, DUD, PTSD, and GAD—suggest potential shared neurobiological substrates. The amygdala-prefrontal circuitry, central to threat detection and impulse control, is implicated in both IED and its comorbidities. For instance, hypoactivity in the ventromedial prefrontal cortex, observed in IED, overlaps with neural signatures of impaired fear extinction in PTSD and reward dysregulation in SUDs. These overlaps support a transdiagnostic model where emotion regulation deficits act as a “final common pathway” for comorbid psychopathology. Such a model aligns with the Research Domain Criteria framework, urging a shift from categorical diagnoses to dimensional constructs like “negative valence systems.”

Notably, the bidirectional correlations between comorbidities (e.g., PTSD-DUD: r = 0.859) may suggest a synergistic “stress-addiction cycle.” Trauma-related hyperarousal may drive substance use as self-medication, while intoxication exacerbates impulsive aggression—a feedback loop perpetuating both disorders. Conversely, inverse correlations (e.g., MDD-Panic: r = - 0.586) hint at protective mechanisms requiring exploration, such as emotional numbing in depression buffering panic symptoms.

### Heterogeneity and Subgroup Differences

Subgroup analyses highlighted that comorbidity risks were systematically higher in community samples compared to clinical populations, suggesting that selection biases in treatment-seeking cohorts may obscure the true syndromic burden of IED. This differential also may reflect cultural and systemic differences in emotional expression and help-seeking behaviors, which warrants further investigation. Diagnostic criteria (DSM-IV vs. DSM-5) did not significantly moderate outcomes, suggesting that evolving definitions have not substantially altered the fundamental comorbidity profile of IED, though differences in symptom thresholds could affect prevalence in subtler ways.

### Limitations and Future Directions

While this meta-analysis offers a comprehensive synthesis, several limitations must be acknowledged. The extreme heterogeneity observed across studies suggests substantial variation in study designs, sample characteristics, and assessment methodologies. Although Bayesian modeling and sensitivity analyses attenuated some concerns, future research should prioritise longitudinal, cross-cultural investigations to disentangle causal pathways and assess how sociodemographic variables, trauma exposure, and early conduct problems moderate the development of IED and its comorbidities. None-reported moderators

## Conclusion

This research reinforces the view of IED as a complex, highly comorbid disorder that bridges traditional diagnostic boundaries. Despite its substantial individual and societal burden, IED remains notably absent from global mental health strategies. Given its strong associations with violence, self-harm, and substance misuse, neglecting IED in public health planning represents a missed opportunity for early intervention and risk reduction. Targeted screening initiatives, particularly among adolescents and young adults with behavioral problems, could facilitate earlier identification and intervention. Furthermore, given the high co-occurrence with mood and anxiety disorders, integrated care models that combine emotional regulation therapies with substance use interventions are urgently needed.

## Supporting information

Appendix I Search terms

Appendix II Data Extraction

Appendix III ROB Results

## Data Availability

All data produced in the present study are available upon reasonable request to the authors.

---

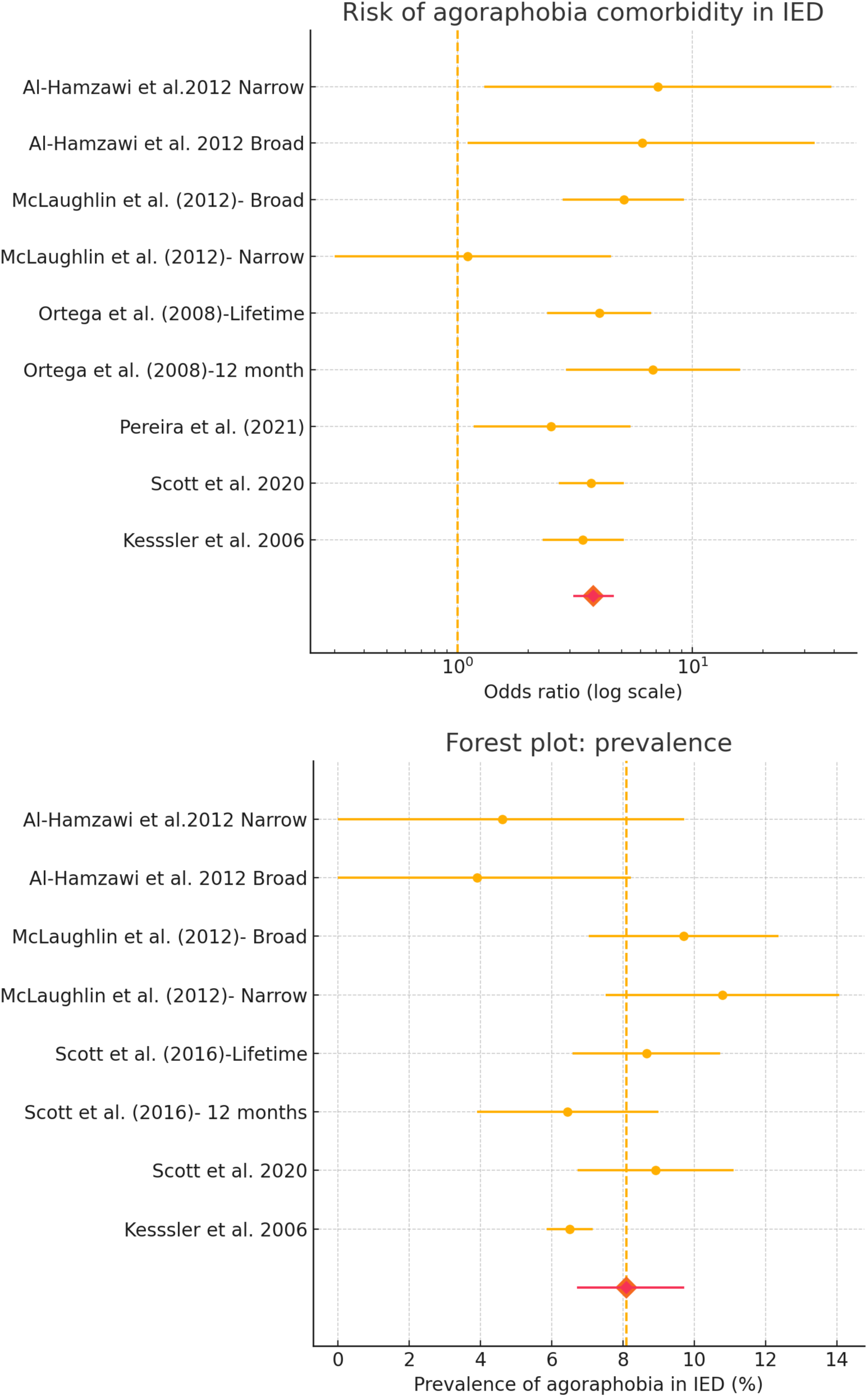
**Agoraphobia Risk and Prevalence Forest Plots**

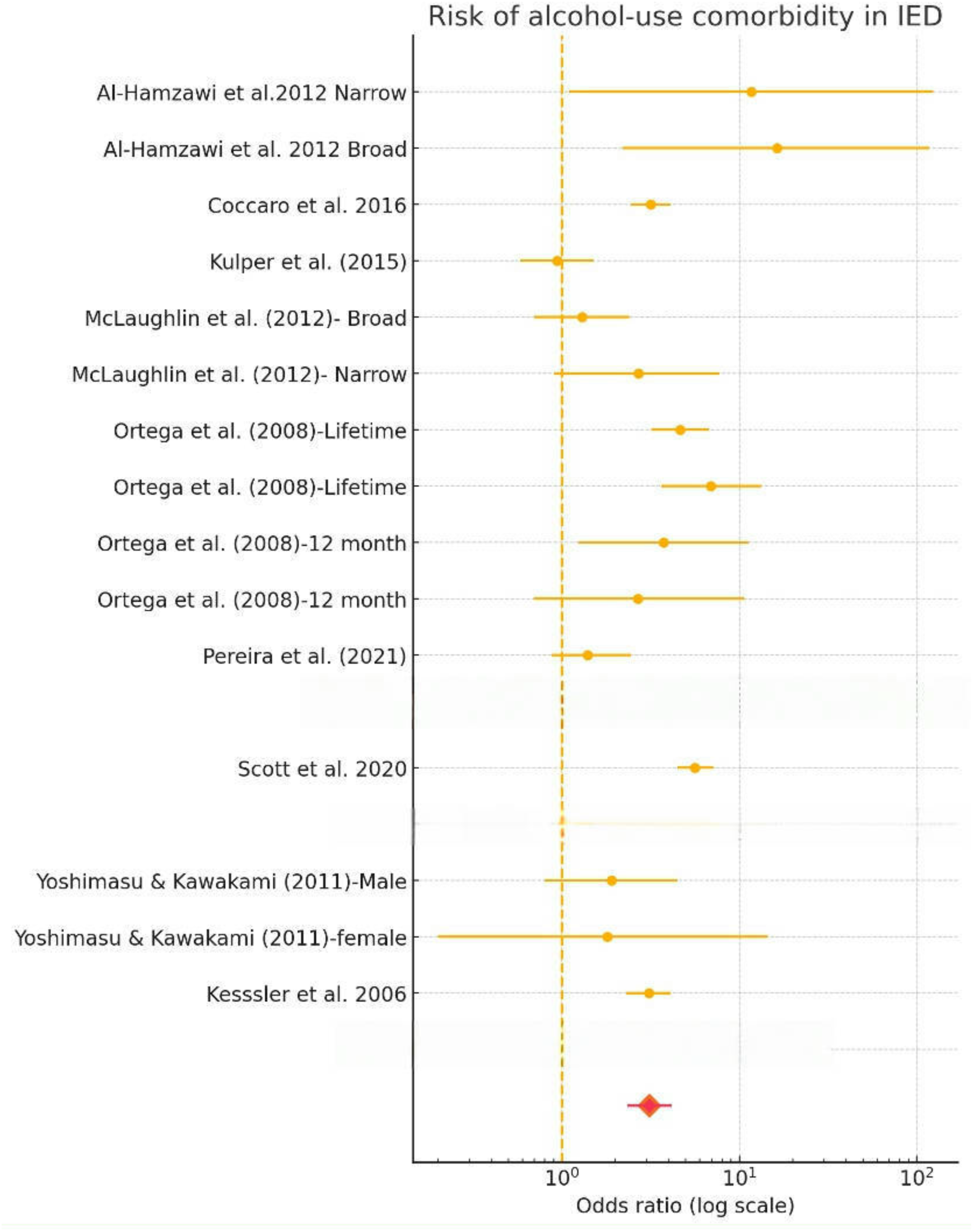
**AUD Risk Forest Plot**

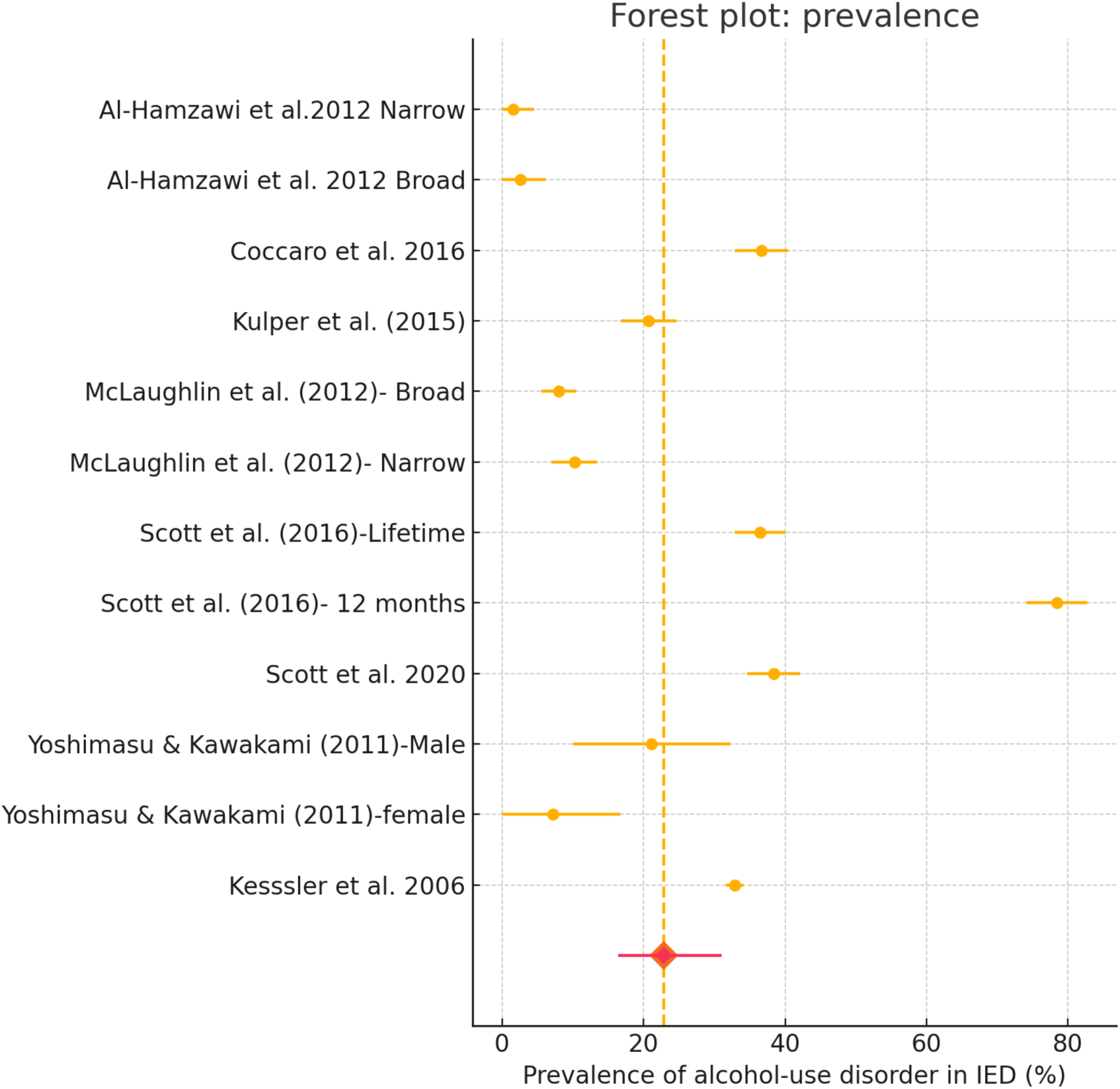
**AUD Prevalence Forest Plot**

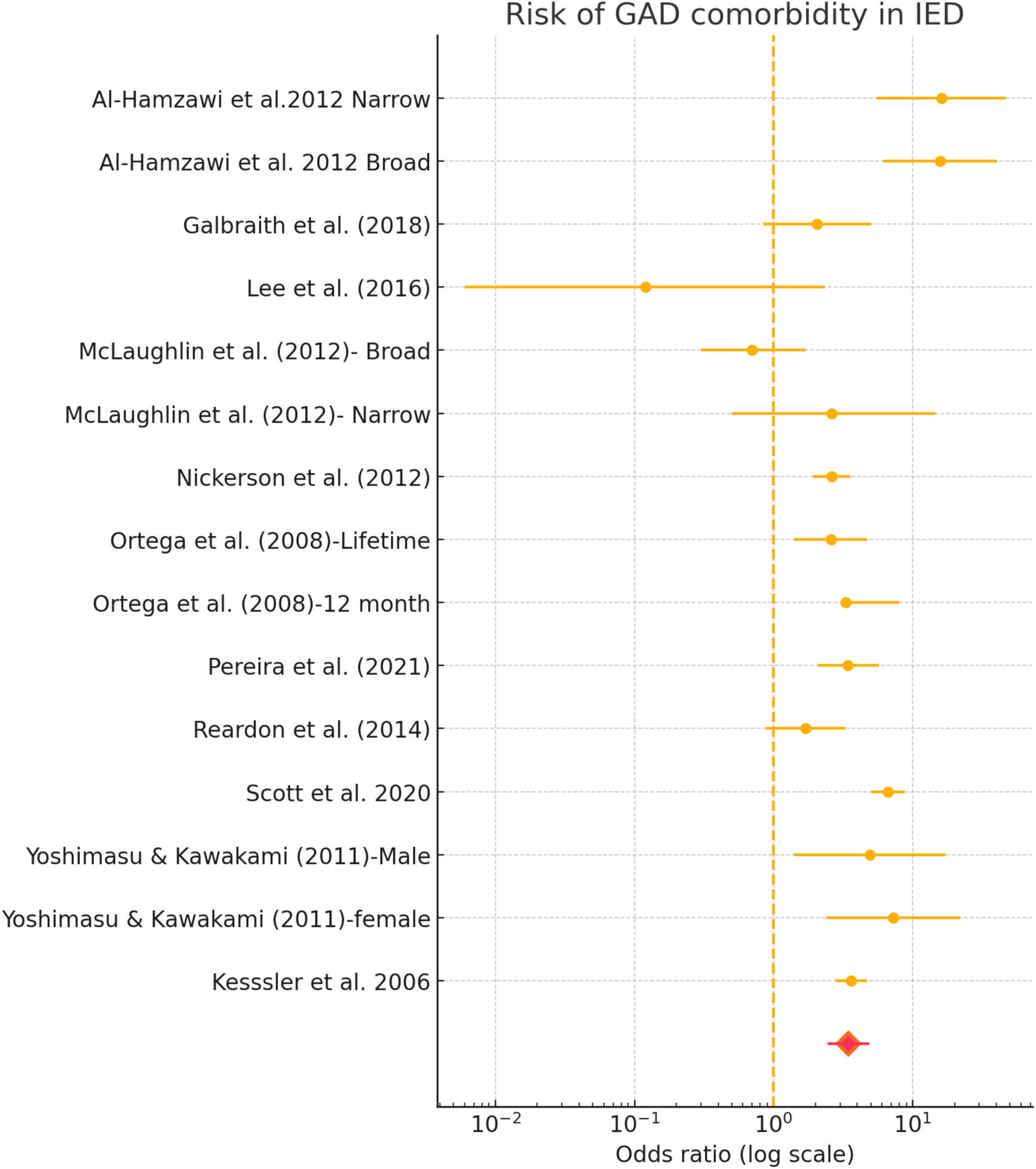
**GAD Risk Forest Plot**

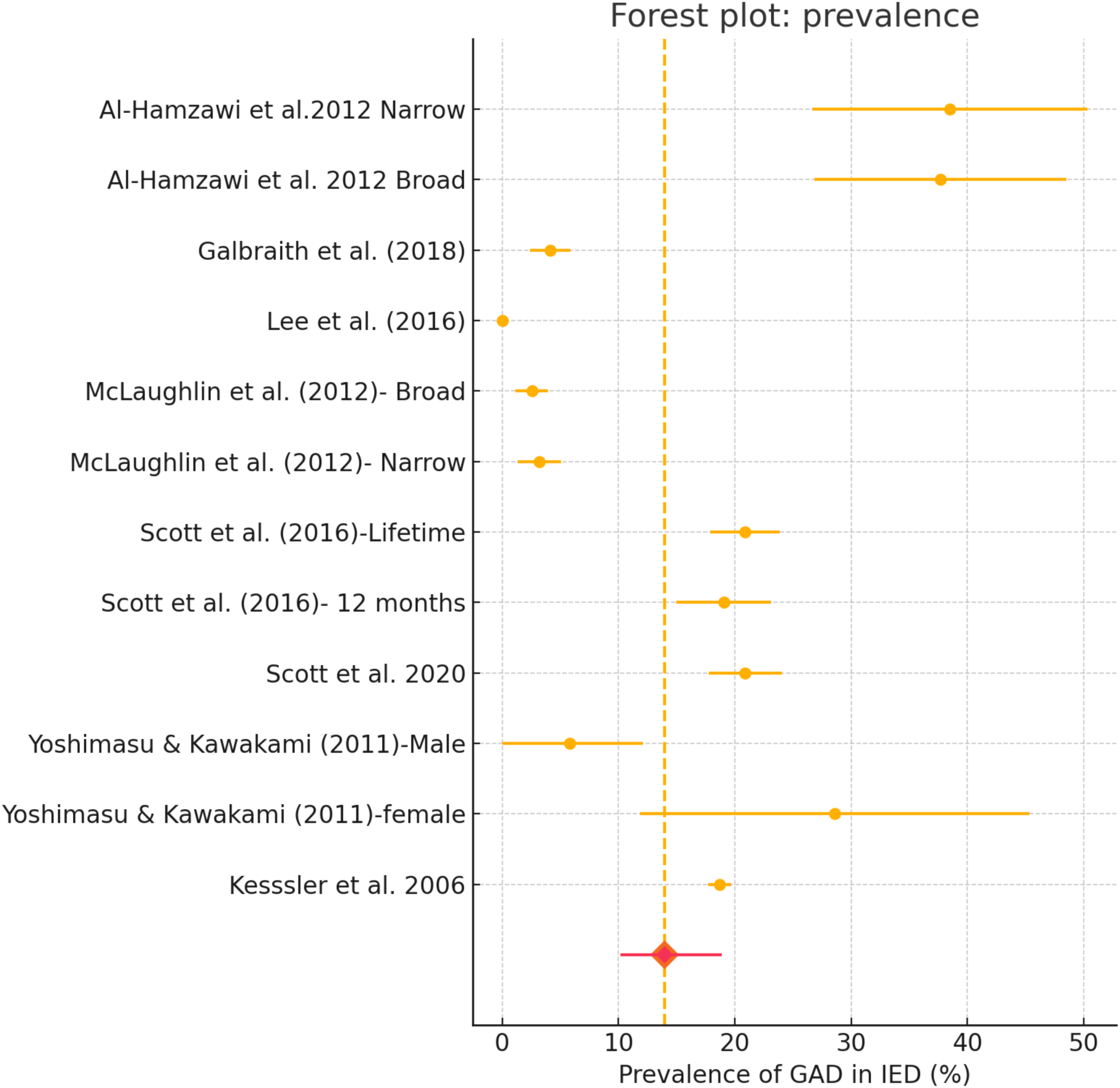
**GAD Prevalence Forest Plot**

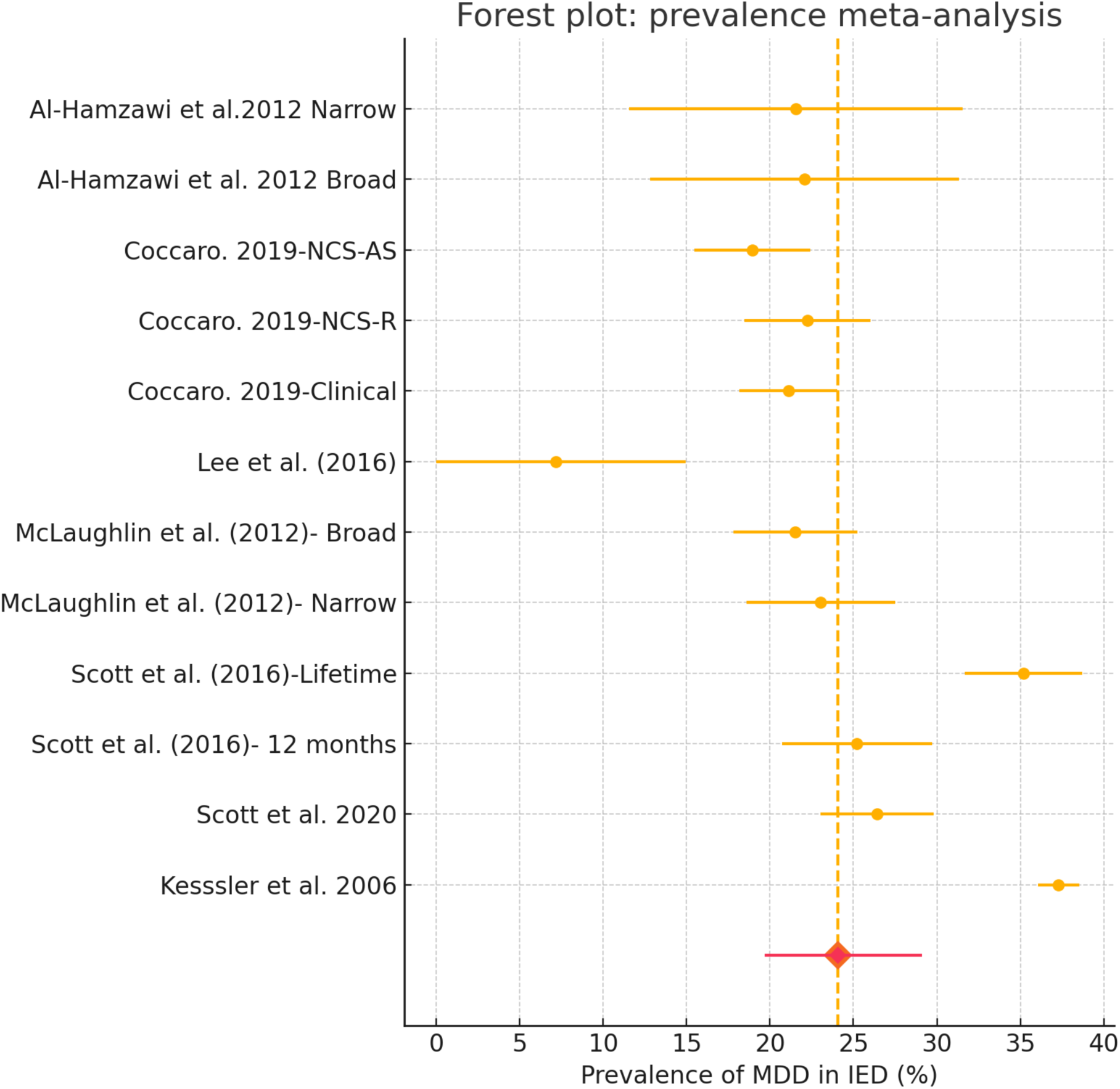
**MDD Prevalence Forest Plot**

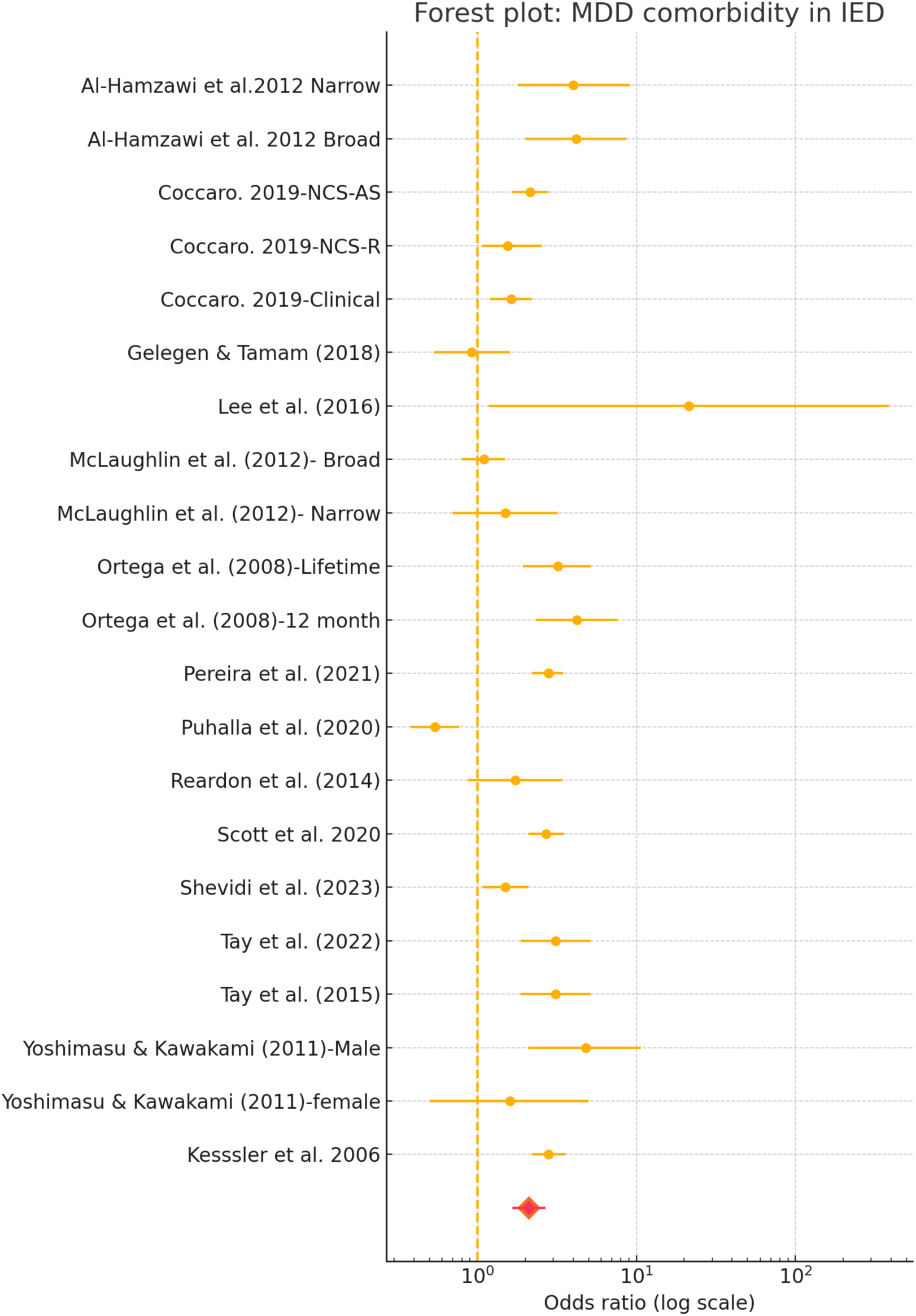
**MDD Risk Forest Plot**

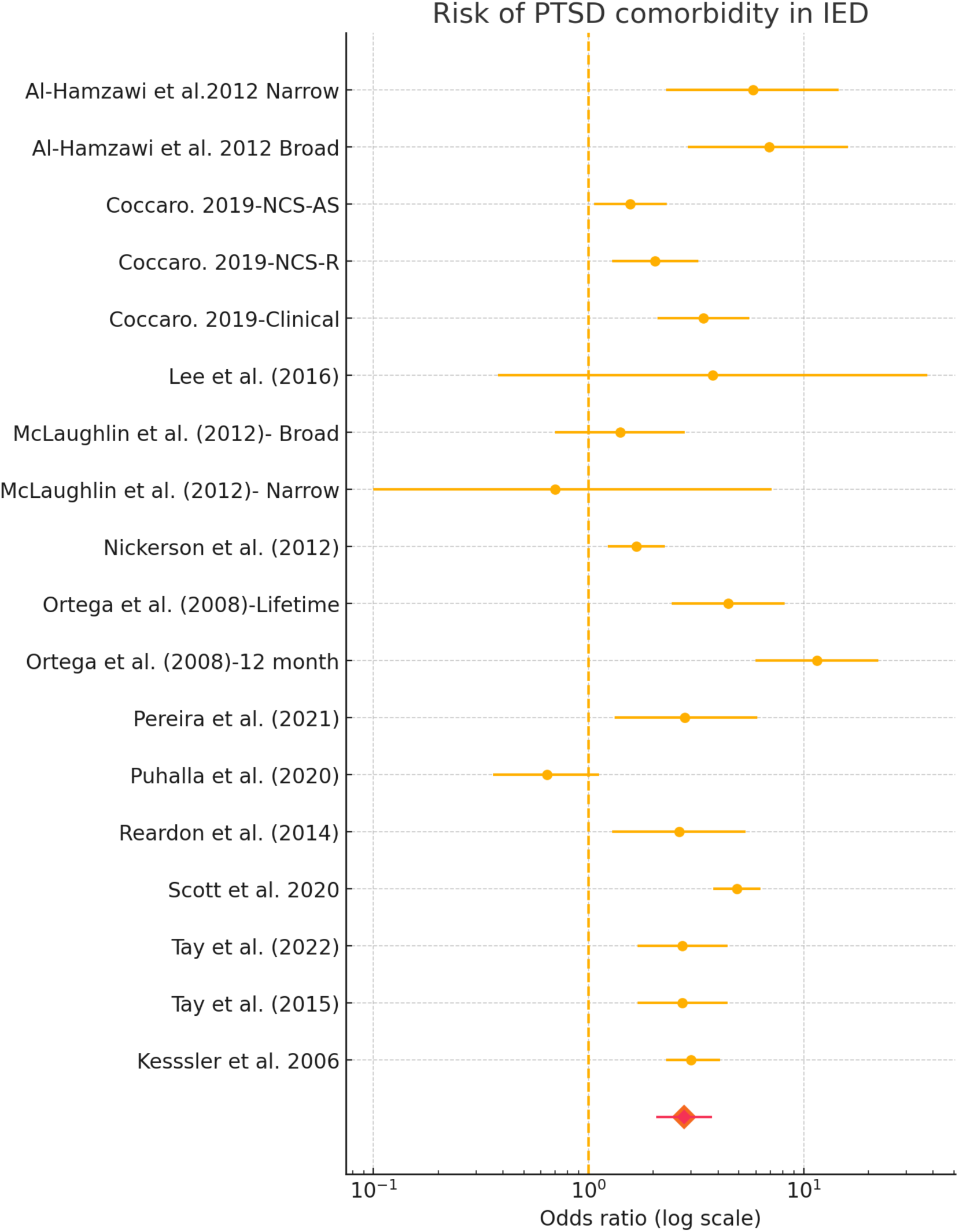
**PTSD Risk Forest Plot**

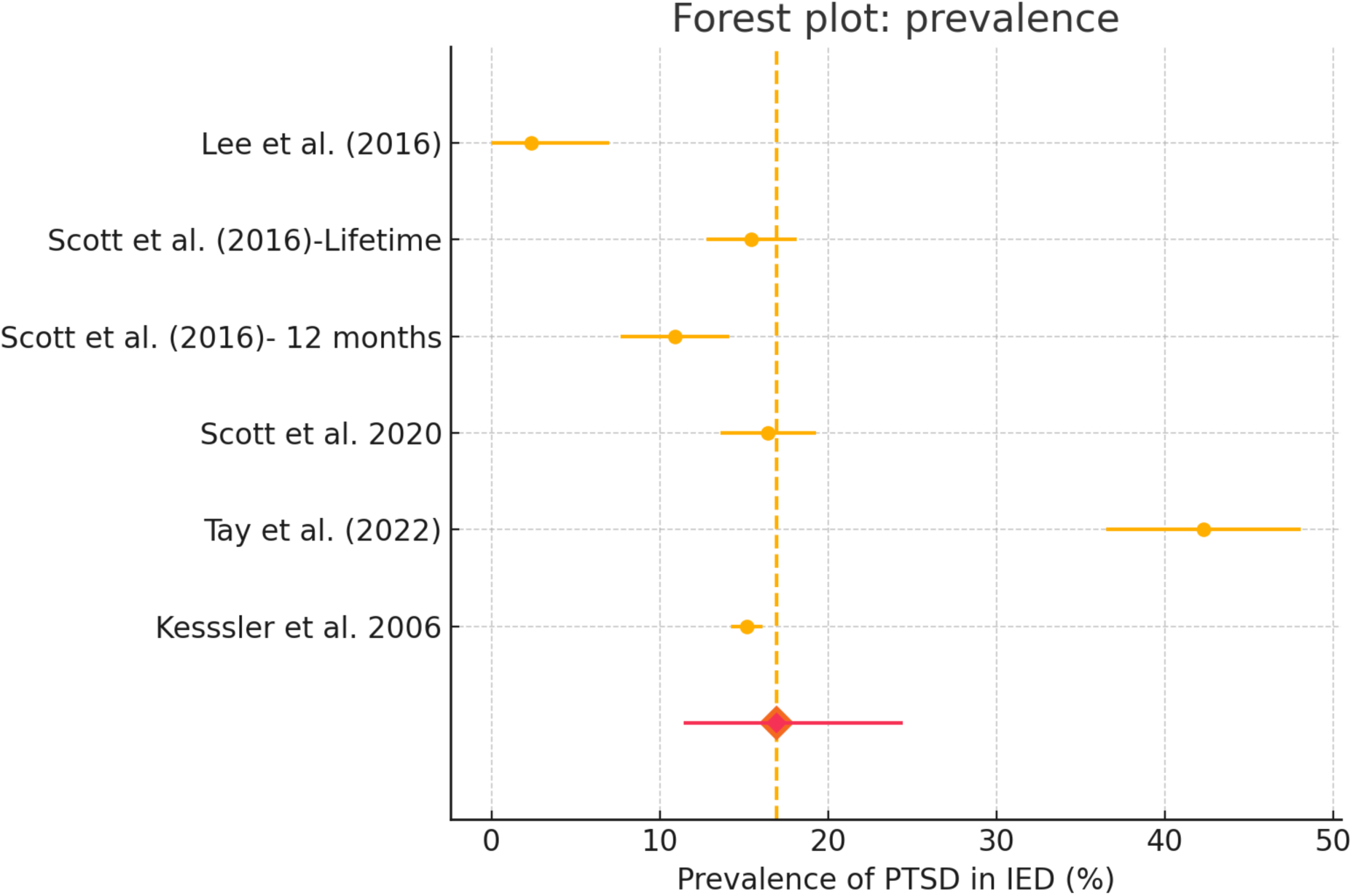
**PTSD Prevalence Forest Plot**

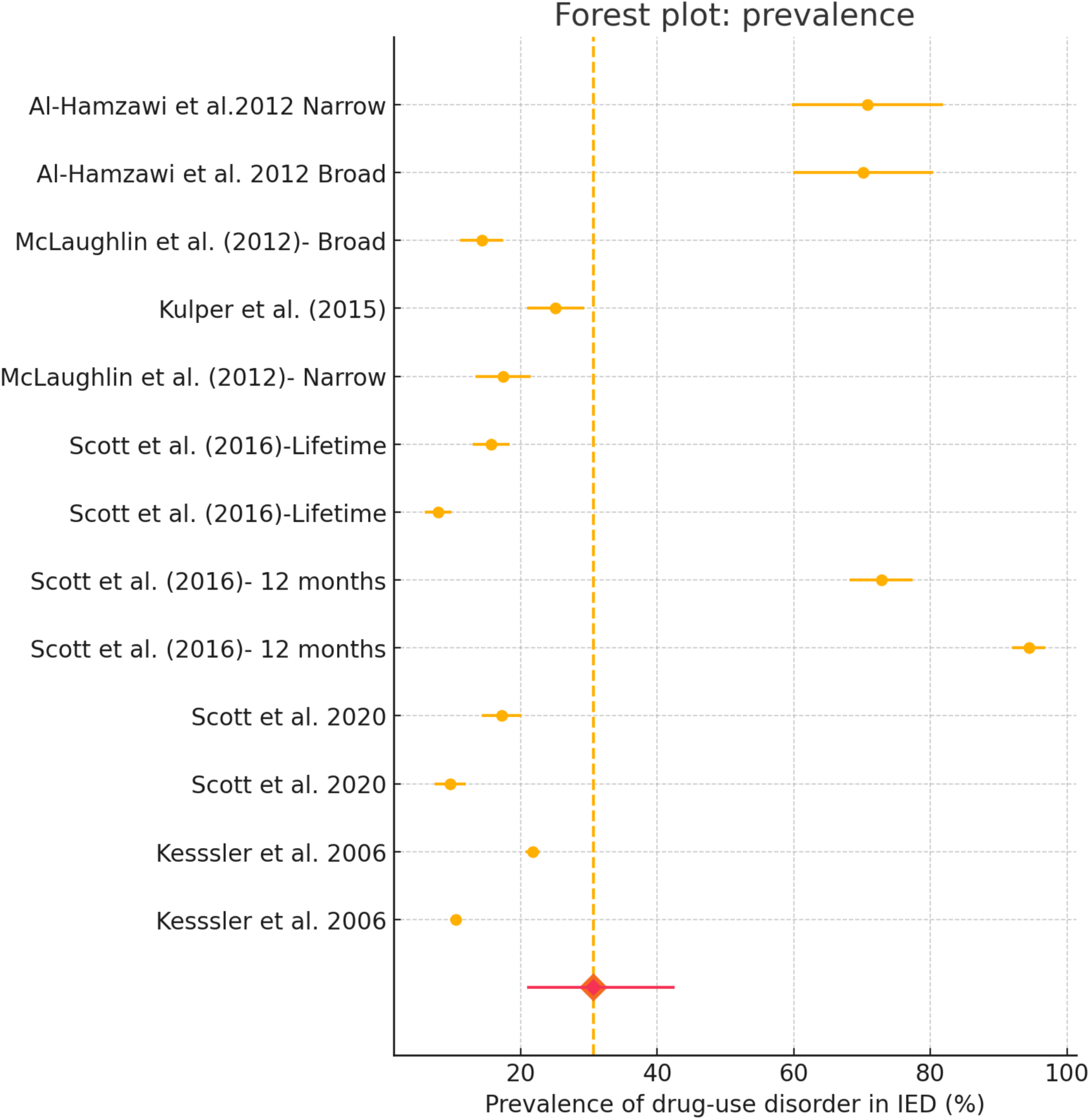
**DUD Prevalence Forest Plot**

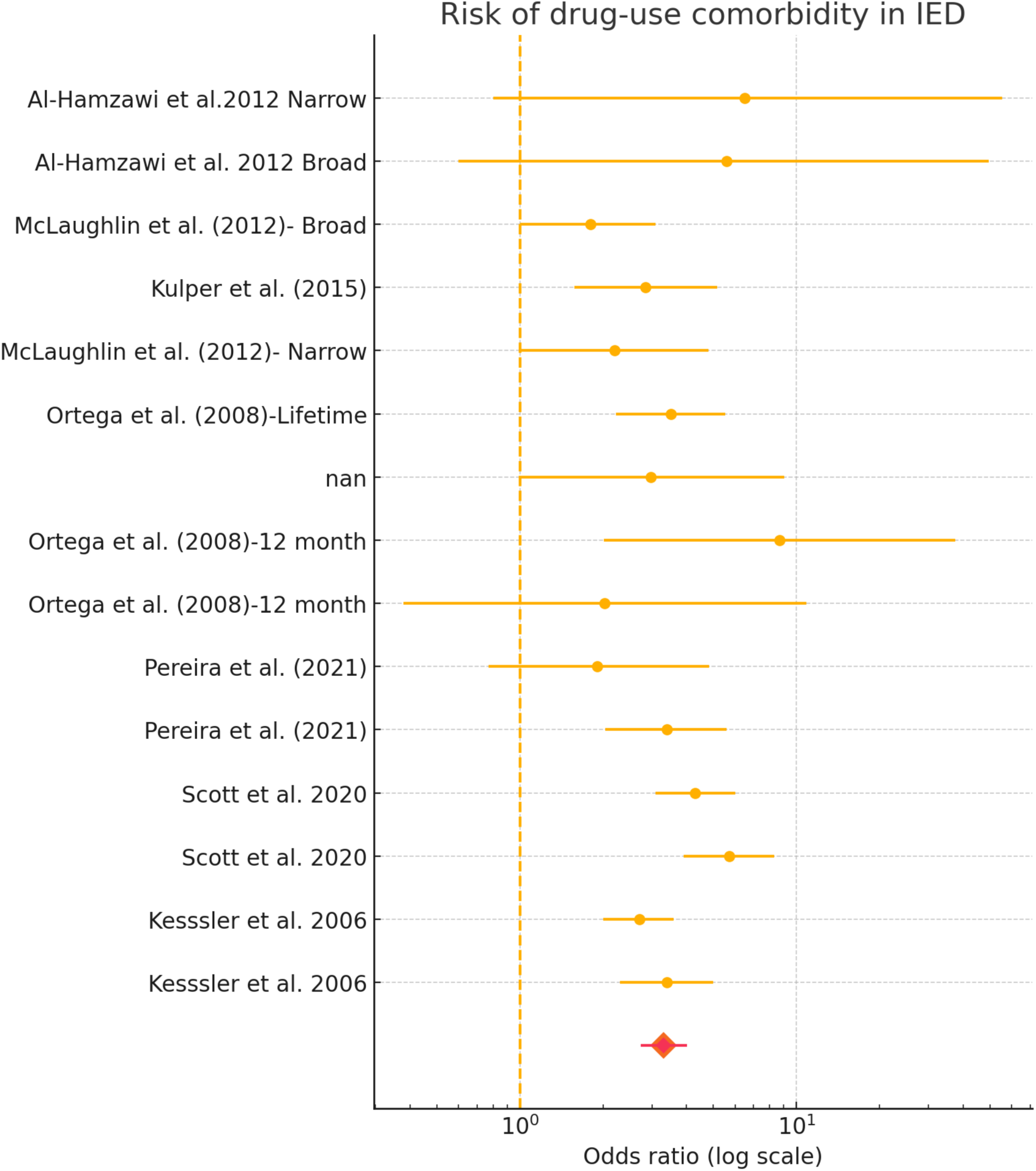
**DUD Risk Forest Plot**

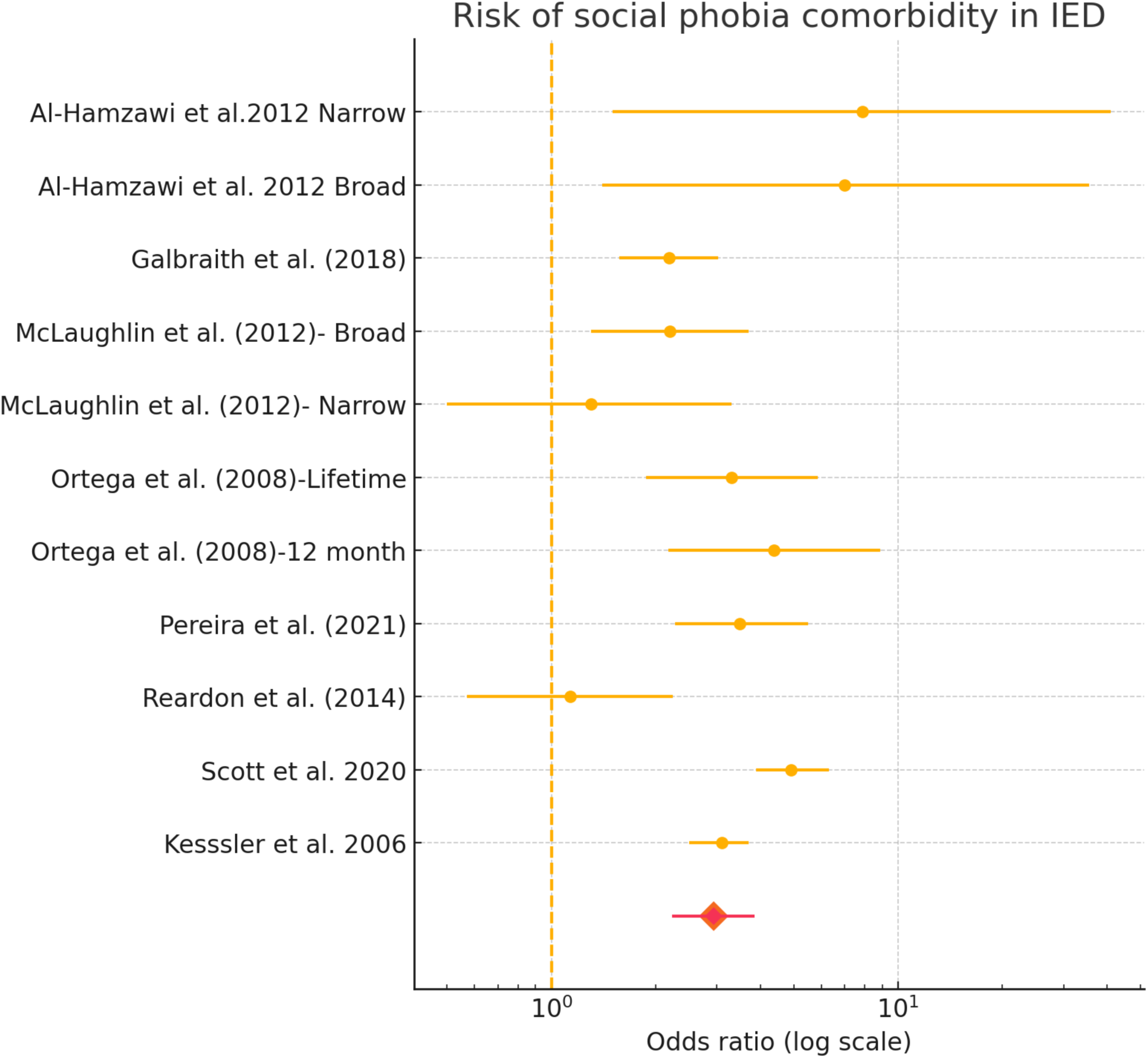
**Social Phobia Forest Plot**

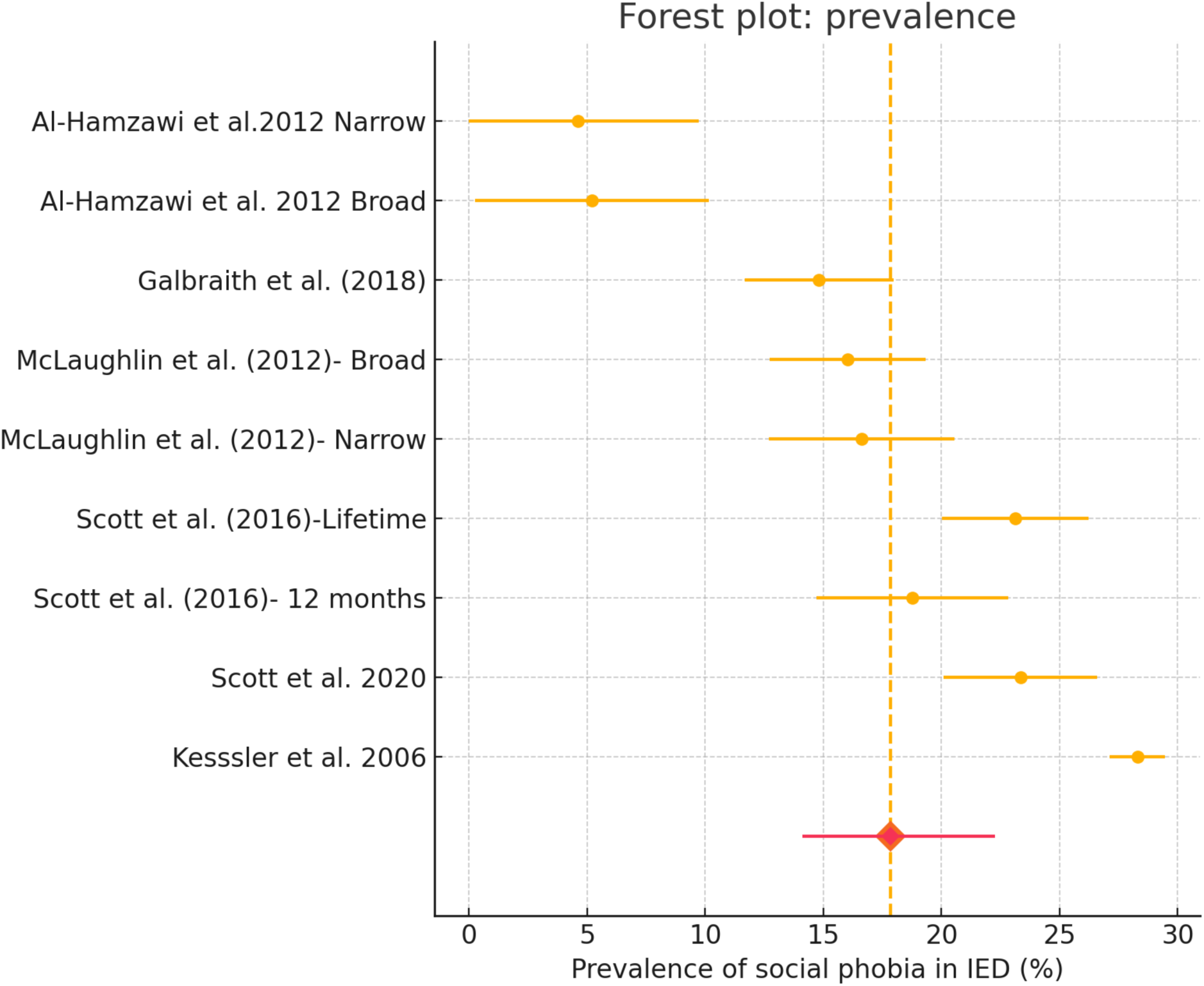
**Social Phobia Prevalence Forest Plot**

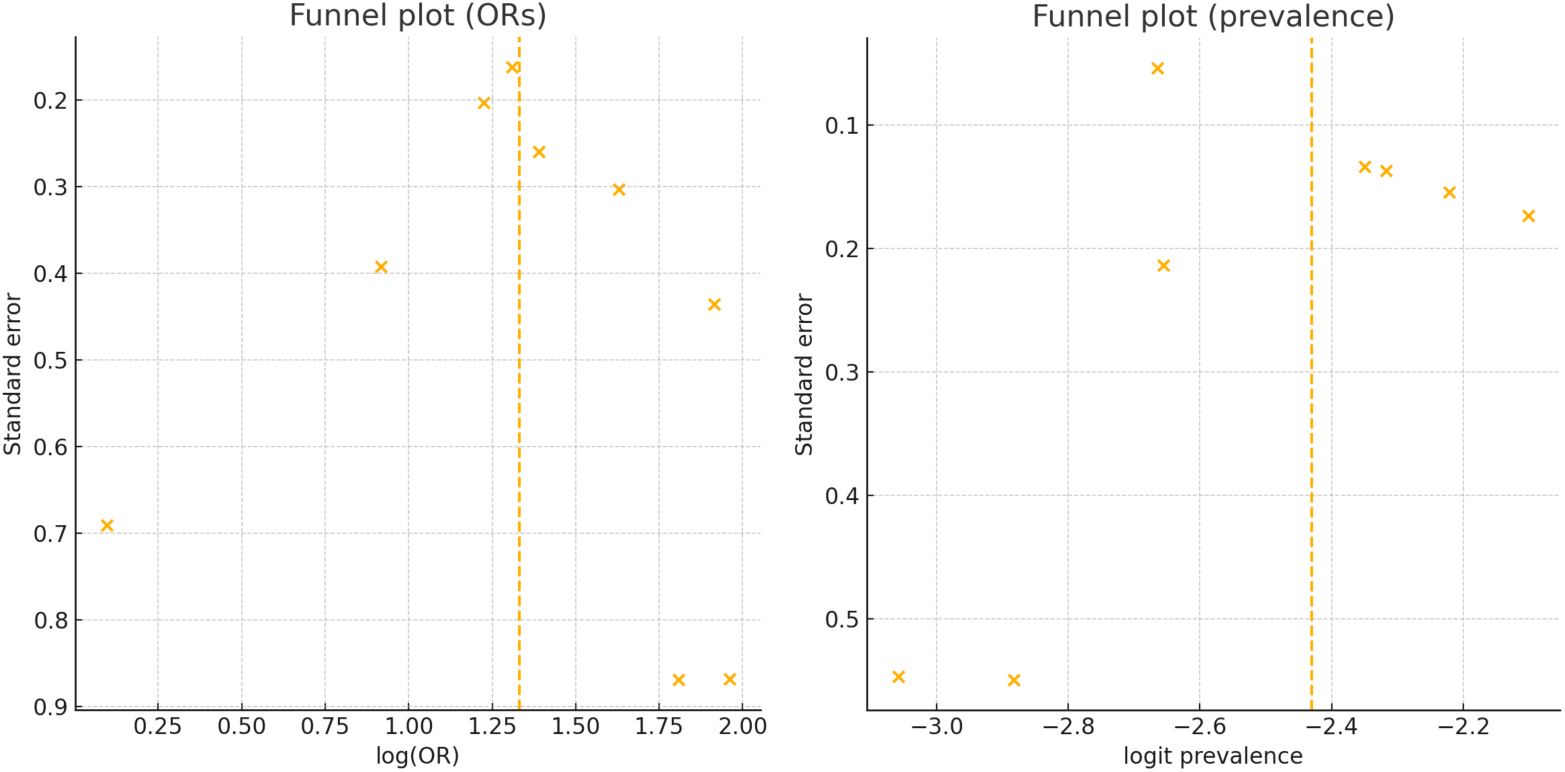
**Agoraphobia Funnel Plots**

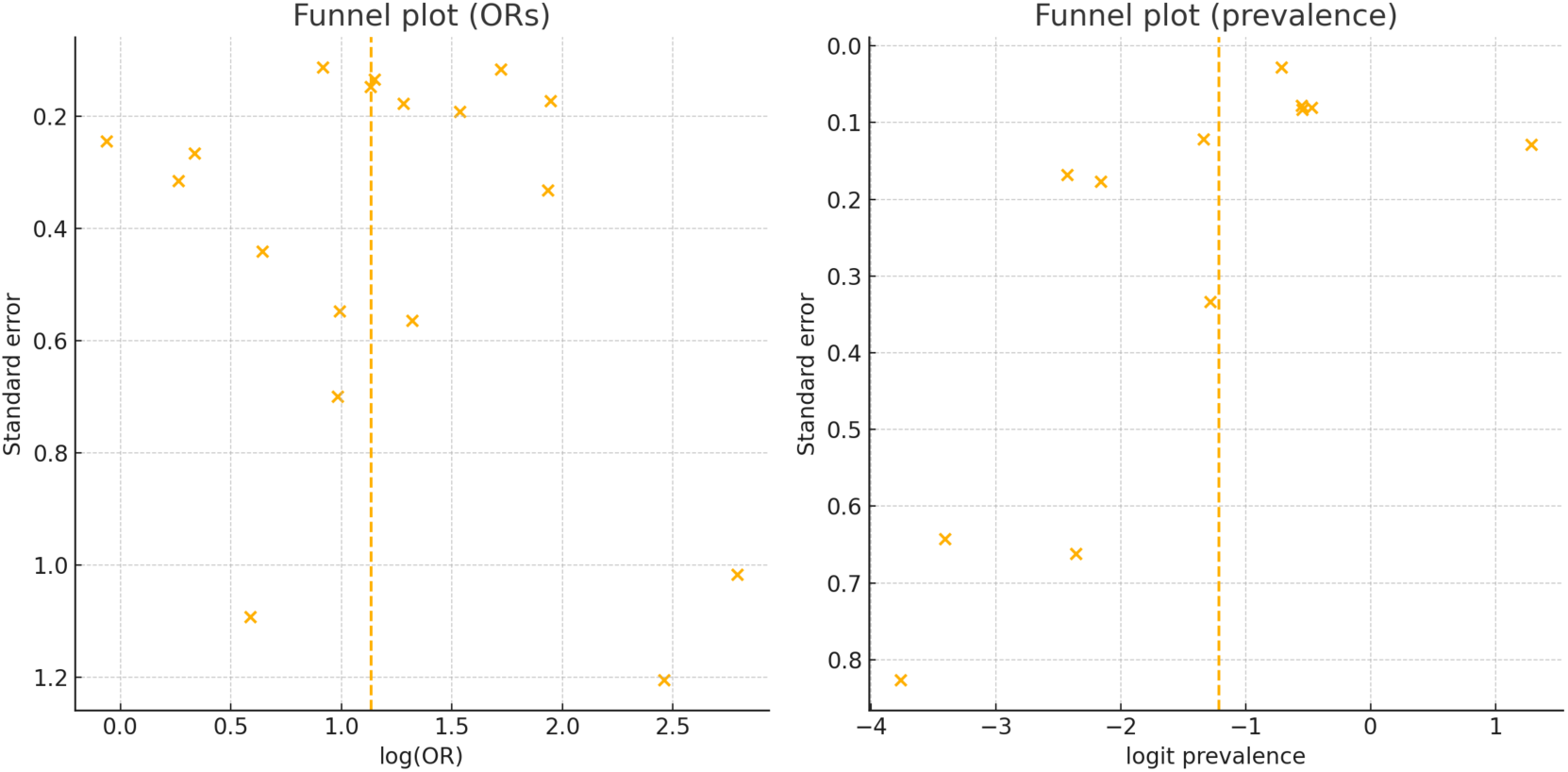
**AUD Funnel Plots**

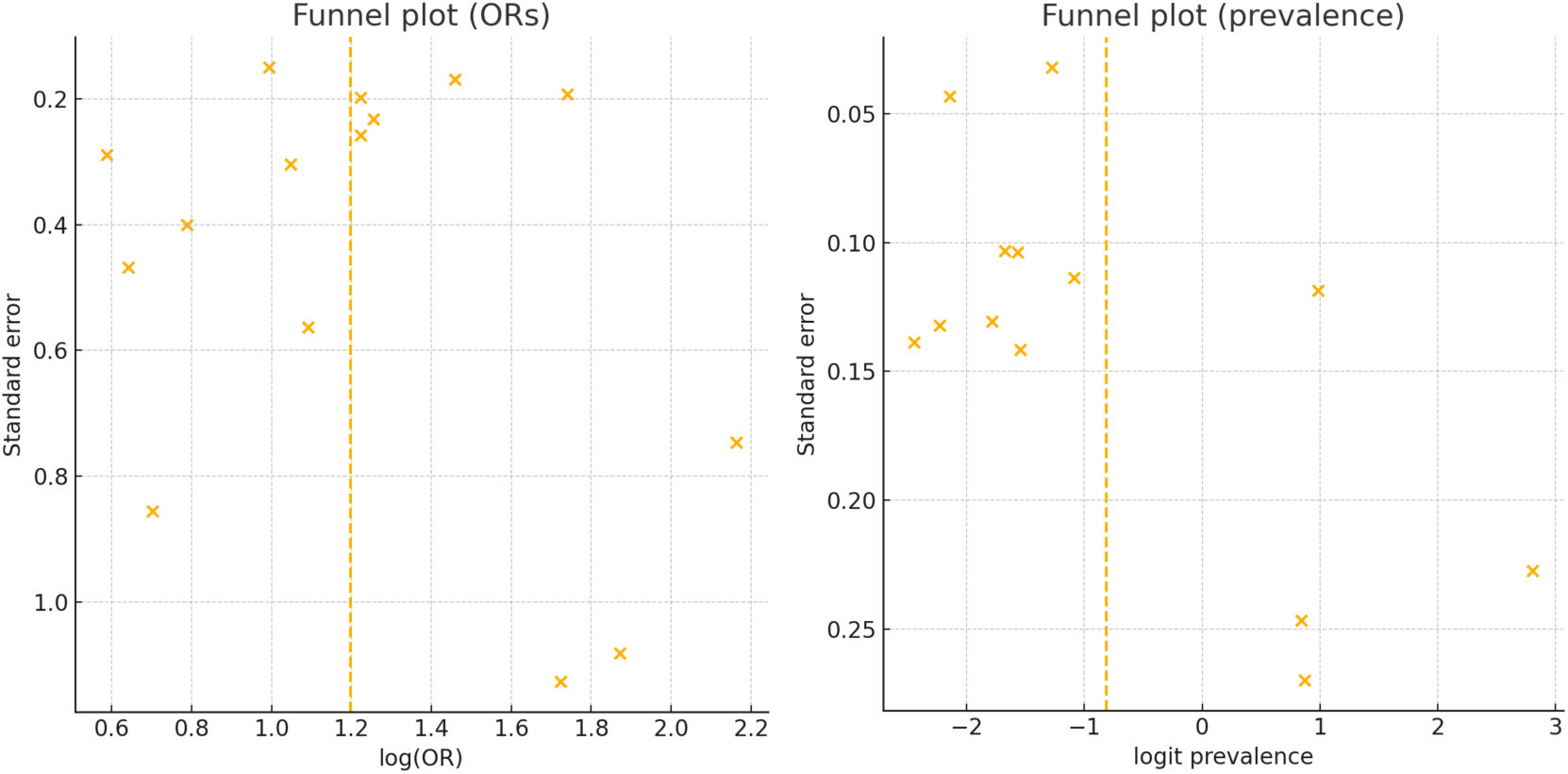
**DUD Funnel Plots**

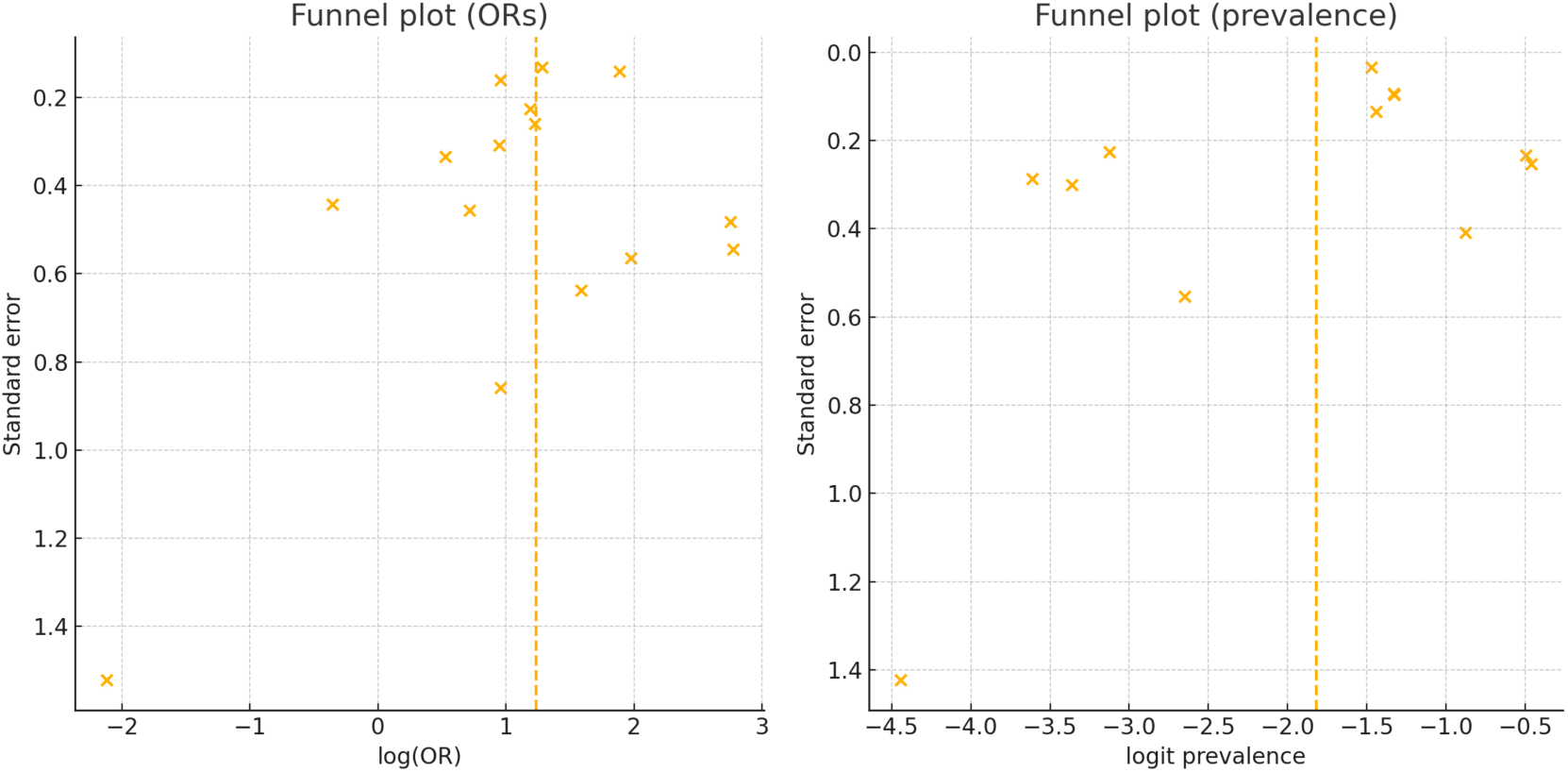
**GAD Funnel Plots**

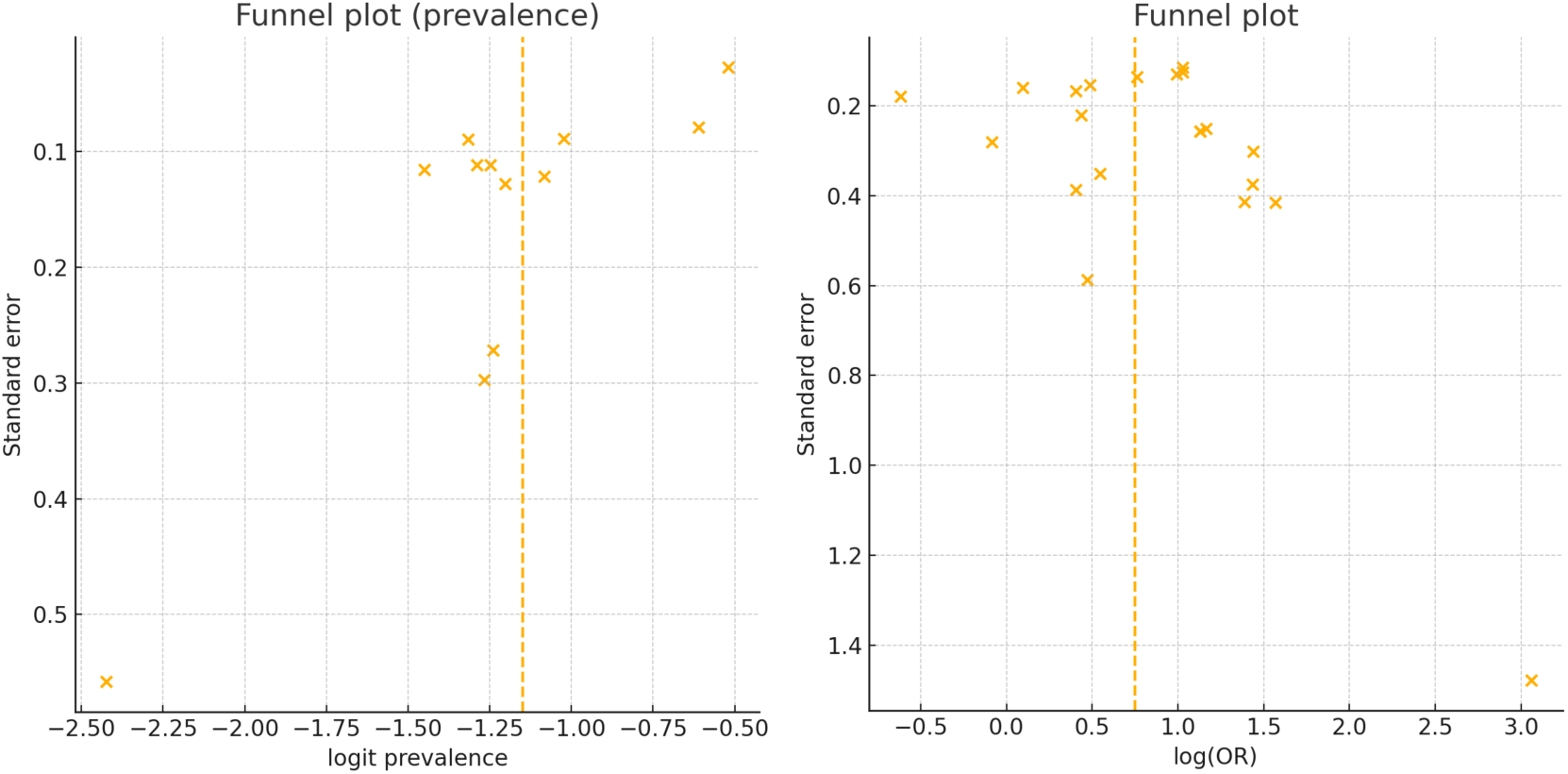
**MDD Funnel Plots**

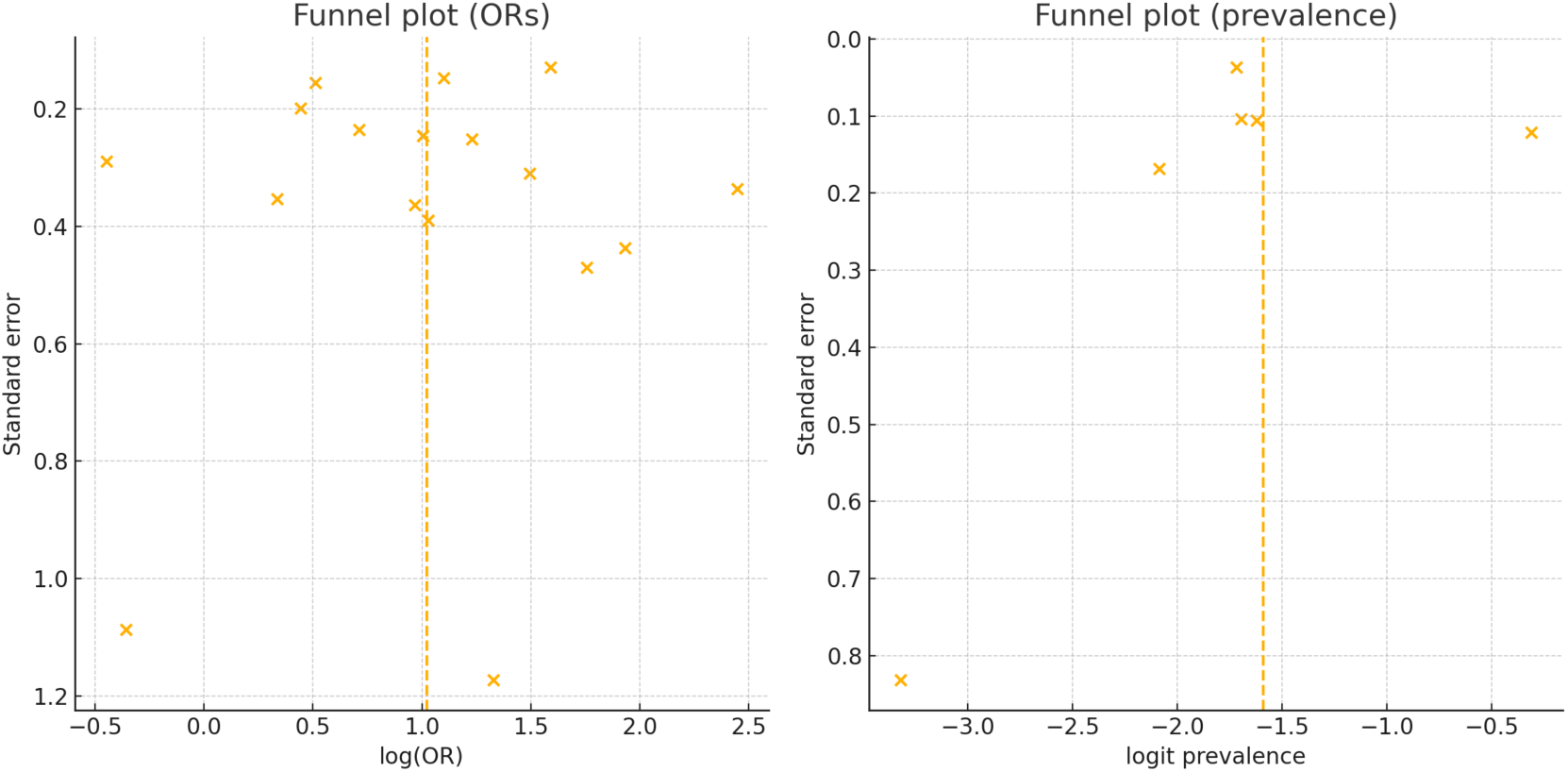
**PTSD Funnel Plots**

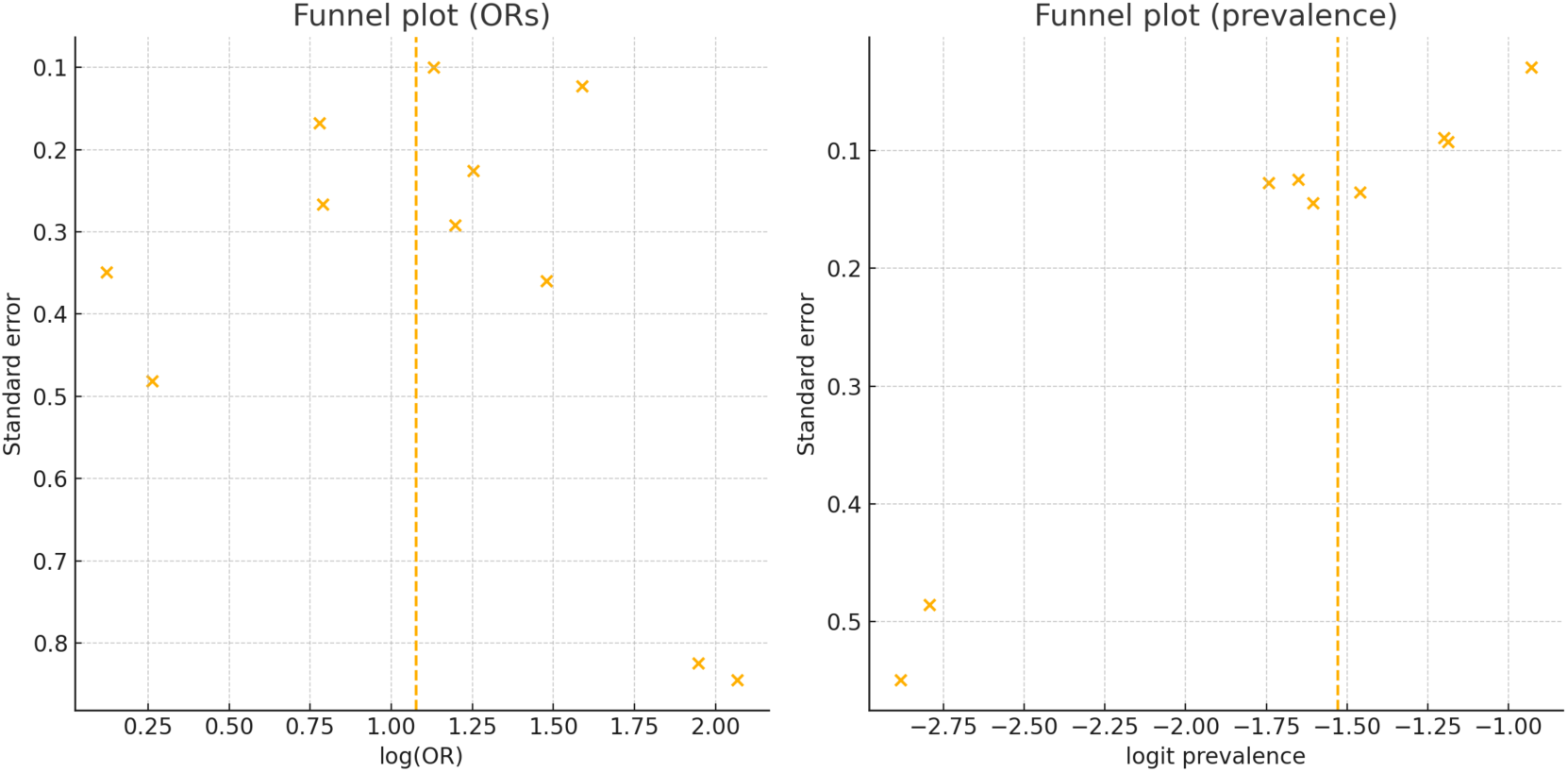
**Social Phobia Funnel Plots**

