## Appendix I Search terms for "Hidden Storms Within: Prevalence and Risk of Comorbidity in Intermittent Explosive Disorder: A Systematic Review and Bayesian Multilevel Meta-Analysis"

**Appendix I. Search terms categorised by concepts and database adaptions**

**Concept 1: Intermittent Explosive Disorder**

("Intermittent Explosive Disorder" OR IED OR

"Explosive Behavior" OR "Aggressive Outbursts" OR

"Impulsive Aggression" OR "Rage Disorder" OR

"Episodic Dyscontrol Syndrome" OR "Anger Attacks")

**Concept 2: Comorbidity & Co-occurrence**

(Comorbid* OR "Co-occurrence" OR "Co-morbidity" OR

"Coexisting" OR "Concurrent" OR "Dual Diagnosis" OR

"Multiple Disorders" OR "Associated Disorders" OR

"Psychiatric Comorbidity" OR "Mental Health Comorbidity")

**Concept 3: Epidemiological Measures**

(Prevalence OR Incidence OR Risk OR "Odds Ratio" OR

"Relative Risk" OR "Risk Factors" OR Epidemiology OR

"Cross-Sectional" OR "Lifetime Prevalence" OR

"12-Month Prevalence" OR Frequency OR Rate)

**Database-Specific Adaptations**

**PubMed/MEDLINE**

#1 MeSH:

("Intermittent Explosive Disorder"[Mesh] OR

"Aggression"[Mesh] OR "Impulsive Behavior"[Mesh])

#2 Free-text:

("Intermittent Explosive Disorder" OR IED OR "Rage Disorder")

#3 MeSH:

("Comorbidity"[Mesh] OR "Dual Diagnosis"[Mesh])

#4 Free-text:

(Comorbid* OR "Co-occurrence" OR "Multiple Disorders")

#5 MeSH:

("Prevalence"[Mesh] OR "Epidemiology"[Mesh] OR "Risk"[Mesh])

#6 Free-text:

(Prevalence OR "Odds Ratio" OR "Risk Factors")

#7 Combine:

(#1 OR #2) AND (#3 OR #4) AND (#5 OR #6)

Filters: Humans, 1980-2025

**PsycINFO (Ovid)**

1. exp Intermittent Explosive Disorder/

2. (IED or "intermittent explosive").ti,ab,kf.

3. 1 or 2

4. exp Comorbidity/

5. (comorbid* or co-occur* or "dual diagnosis").ti,ab,kf.

6. 4 or 5

7. exp Prevalence/ or exp Risk Factors/

8. (prevalence or risk or epidemiol*).ti,ab,kf.

9. 7 or 8

10. 3 and 6 and 9

**EMBASE**

[intermittent explosive disorder]/de OR

(ied OR "aggressive outbursts" OR "rage disorder")

AND

[comorbidity]/de OR (comorbid* OR "co-occurrence")

AND

[prevalence]/de OR [risk]/de OR

(prevalence OR "odds ratio" OR epidemiology)

LIMIT: [1980-2024] AND [human]
