## Appendix II Data Extraction for "Hidden Storms Within: Prevalence and Risk of Comorbidity in Intermittent Explosive Disorder: A Systematic Review and Bayesian Multilevel Meta-Analysis"

| **Study ID** | **Study Year** | **Sample Size** | **Sample Type** | **Study Design** | **Diagnostic Criteria** | **(Specific Comorbidity / Correlate)** | **pre% (standard error)** | **OR** | **95% CI** |  |  |  |
| --- | --- | --- | --- | --- | --- | --- | --- | --- | --- | --- | --- | --- |
| Al-Hamzawi et al.2012 Narrow | 2012 | 65 | Community | Cross-sectional survey | DSM IV | Agoraphobia | 4.3 (3.3) | 7.1 | 1.3-39.1 |  |  |  |
| Al-Hamzawi et al. 2012 Broad | 2012 | 77 | Community | Cross-sectional survey | DSM IV | Agoraphobia | 3.9 (3.0) | 6.1 | 1.1-33.1 |  |  |  |
| McLaughlin et al. (2012)- Broad | USA | 474 | Community | Cross-sectional survey | DSM IV | Agoraphobia | 9.7(2.6) | 5.1 | 2.8-9.2 |  |  |  |
| McLaughlin et al. (2012)- Narrow | USA | 343 | Community | Cross-sectional survey | DSM IV | Agoraphobia | 10.7(4.1) | 1.1 | 0.3-4.5 |  |  |  |
| Ortega et al. (2008)-Lifetime | USA | 158 | Community | Cross-sectional survey | DSM IV | Agoraphobia |  | 4.01 | 2.41-6.68 |  |  |  |
| Ortega et al. (2008)-12 month | Mexico | 106 | Community | Cross-sectional survey | DSM IV | Agoraphobia |  | 6.78 | 2.89-15.94 |  |  |  |
| Pereira et al. (2021) | Brazil | 171 | Community | Cross-sectional survey | DSM 5 | Agoraphobia |  | 2.5 | 1.17-5.44 |  |  |  |
| Scott et al. (2016)-Lifetime | Cross-national (WMH) | 705 | Community | Cross-sectional study | DSM 5 | Agoraphobia | 8.7 (1.2) |  |  |  |  |  |
| Scott et al. (2016)- 12 months | Cross-national (WMH) | 357 | Community | Cross-sectional study | DSM 5 | Agoraphobia | 6.5 (1.5) |  |  |  |  |  |
| Scott et al. 2020 | Cross-national (WMH) | 651 | Community | Cross-sectional study | DSM 5 | Agoraphobia | 8.9(1.2) | 3.7 | 2.7-5.1 |  |  |  |
| Kesssler et al. 2006 | USA | 5692 | Community | Cross-sectional study | DSM IV | Agoraphobia | 6.5 (1.1) | 3.4 | 2.3-5.1 |  |  |  |
| **Study ID** | **Study Year** | **Sample Size** | **Sample Type** | **Study Design** | **Diagnostic Criteria** | **(Specific Comorbidity / Correlate)** | **pre% (standard error)** | **OR** | **95% CI** | **p-value** |  |  |
| Al-Hamzawi et al.2012 Narrow | 2012 | 65 | Community | Cross-sectional survey | DSM IV | Alcohol dependence with abuse | 2.2 (2.2) | 11.7 | 1.1-123.6 | 0.04 |  |  |
| Al-Hamzawi et al. 2012 Broad | 2012 | 77 | Community | Cross-sectional survey | DSM IV | Alcohol dependence with abuse | 2.8 (2.1) | 16.3 | 2.2-118.4 | 0.005 |  |  |
| Coccaro et al. 2016 | 2016 | 619 | Clinical | Cross-sectional survey | DSM 5 | Alcohol use disorder | 36.60% | 3.16 | 2.43-4.11 |  |  |  |
| Kulper et al. (2015) | 2015 | 410 | Clinical | Cross-sectional survey | DSM 5 | Alcohol dependence | 20.8 vs 21.8 | 0.94 | 0.58-1.51 | < 0.01 |  |  |
| McLaughlin et al. (2012)- Broad | USA | 474 | Community | Cross-sectional survey | DSM IV | Alcohol dependence with abuse | 8.1 (1.7) | 1.3 | 0.7-2.4 |  |  |  |
| McLaughlin et al. (2012)- Narrow | USA | 343 | Community | Cross-sectional survey | DSM IV | Alcohol dependence with abuse | 10.2 (2.5) | 2.7 | 0.9-7.7 |  |  |  |
| Ortega et al. (2008)-Lifetime | USA | 158 | Community | Cross-sectional survey | DSM IV | Alcohol abuse |  | 4.65 | 3.20-6.76 |  |  |  |
| Ortega et al. (2008)-Lifetime | USA |  | Community | Cross-sectional survey | DSM IV | Alcohol dependence |  | 6.91 | 3.61-13.22 |  |  |  |
| Ortega et al. (2008)-12 month | Mexico | 106 | Community | Cross-sectional survey | DSM IV | Alcohol abuse |  | 3.75 | 1.24-11.31 |  |  |  |
| Ortega et al. (2008)-12 month | Mexico |  | Community | Cross-sectional survey | DSM IV | Alcohol dependence |  | 2.67 | 0.69-10.70 |  |  |  |
| Pereira et al. (2021) | Brazil | 171 | Community | Cross-sectional survey | DSM 5 | Alcohol dependence |  | 1.4 | 0.87-2.46 |  |  |  |
|  |  |  |  |  |  | Alcohol abuse |  | 2.5 | 2.02-3.14 |  |  |  |
| Scott et al. (2016)-Lifetime | Cross-national (WMH) | 705 | Community | Cross-sectional study | DSM 5 | Alcohol abuse | 36.5 (2.3) |  |  |  |  |  |
|  |  |  |  |  |  | Alcohol dependence | 17.2 (2.0) |  |  |  |  |  |
| Scott et al. (2016)- 12 months | Cross-national (WMH) | 357 | Community | Cross-sectional study | DSM 5 | Alcohol abuse | 78.3 (3.3) |  |  |  |  |  |
|  |  |  |  |  |  | Alcohol dependence | 92.4 (2.7) |  |  |  |  |  |
| Scott et al. 2020 | Cross-national (WMH) | 651 | Community | Cross-sectional study | DSM 5 | Alcohol abuse | 38.4(2.4) | 5.6 | 4.5-7.1 |  |  |  |
|  |  |  |  |  |  | Alcohol dependence | 20.0(2.2) | 7 | 5.0-9.8 |  |  |  |
| Yoshimasu & Kawakami (2011)-Male | Japan | 52 | Community | Cross-sectional study | DSM IV | Alcohol abuse | 22.00% | 1.9 | 0.8-4.5 |  |  |  |
| Yoshimasu & Kawakami (2011)-female | Japan | 28 | Community | Cross-sectional study | DSM IV | Alcohol abuse | 8.4 | 1.8 | 0.2-14.5 |  |  |  |
| Kesssler et al. 2006 | USA | 5692 |  | Cross-sectional study | DSM IV | Alcohol abuse | 32.9 (3.0) | 3.1 | (2.3-4.1) |  |  |  |
|  |  |  |  |  |  | Alcohol dependence | 17.0(2.0) | 3.6 | (2.5-5.0) |  |  |  |
| **Study ID** | **Study Year** | **Sample Size** | **Sample Type** | **Study Design** | **Diagnostic Criteria** | **(Specific Comorbidity / Correlate)** | **pre% (standard error)** | **OR** | **95% CI** |  |  |  |
| Al-Hamzawi et al.2012 Narrow | 2012 | 65 | Community | Cross-sectional survey | DSM IV | Alcohol dependence with abuse | 2.2 (2.2) | 11.7 | 1.1-123.6 |  |  |  |
| Al-Hamzawi et al. 2012 Broad | 2012 | 77 | Community | Cross-sectional survey | DSM IV | Alcohol dependence with abuse | 2.8 (2.1) | 16.3 | 2.2-118.4 |  |  |  |
| Coccaro et al. 2016 | 2016 | 619 | Clinical | Cross-sectional survey | DSM 5 | Alcohol use disorder | 36.60% | 3.16 | 2.43-4.11 |  |  |  |
| Kulper et al. (2015) | 2015 | 410 | Clinical | Cross-sectional survey | DSM 5 | Alcohol dependence | 20.8 vs 21.8 | 0.94 | 0.58-1.51 |  |  |  |
| McLaughlin et al. (2012)- Broad | USA | 474 | Community | Cross-sectional survey | DSM IV | Alcohol dependence with abuse | 8.1 (1.7) | 1.3 | 0.7-2.4 |  |  |  |
| McLaughlin et al. (2012)- Narrow | USA | 343 | Community | Cross-sectional survey | DSM IV | Alcohol dependence with abuse | 10.2 (2.5) | 2.7 | 0.9-7.7 |  |  |  |
| Ortega et al. (2008)-Lifetime | USA | 158 | Community | Cross-sectional survey | DSM IV | Alcohol abuse |  | 4.65 | 3.20-6.76 |  |  |  |
| Ortega et al. (2008)-Lifetime | USA |  | Community | Cross-sectional survey | DSM IV | Alcohol dependence |  | 6.91 | 3.61-13.22 |  |  |  |
| Ortega et al. (2008)-12 month | Mexico | 106 | Community | Cross-sectional survey | DSM IV | Alcohol abuse |  | 3.75 | 1.24-11.31 |  |  |  |
| Ortega et al. (2008)-12 month | Mexico |  | Community | Cross-sectional survey | DSM IV | Alcohol dependence |  | 2.67 | 0.69-10.70 |  |  |  |
| Pereira et al. (2021) | Brazil | 171 | Community | Cross-sectional survey | DSM 5 | Alcohol dependence |  | 1.4 | 0.87-2.46 |  |  |  |
|  |  |  |  |  |  | Alcohol abuse |  | 2.5 | 2.02-3.14 |  |  |  |
| Scott et al. (2016)-Lifetime | Cross-national (WMH) | 705 | Community | Cross-sectional study | DSM 5 | Alcohol abuse | 36.5 (2.3) |  |  |  |  |  |
|  |  |  |  |  |  | Alcohol dependence | 17.2 (2.0) |  |  |  |  |  |
| Scott et al. (2016)- 12 months | Cross-national (WMH) | 357 | Community | Cross-sectional study | DSM 5 | Alcohol abuse | 78.3 (3.3) |  |  |  |  |  |
|  |  |  |  |  |  | Alcohol dependence | 92.4 (2.7) |  |  |  |  |  |
| Scott et al. 2020 | Cross-national (WMH) | 651 | Community | Cross-sectional study | DSM 5 | Alcohol abuse | 38.4(2.4) | 5.6 | 4.5-7.1 |  |  |  |
|  |  |  |  |  |  | Alcohol dependence | 20.0(2.2) | 7 | 5.0-9.8 |  |  |  |
| Yoshimasu & Kawakami (2011)-Male | Japan | 52 | Community | Cross-sectional study | DSM IV | Alcohol abuse | 22.00% | 1.9 | 0.8-4.5 |  |  |  |
| Yoshimasu & Kawakami (2011)-female | Japan | 28 | Community | Cross-sectional study | DSM IV | Alcohol abuse | 8.4 | 1.8 | 0.2-14.5 |  |  |  |
| Kesssler et al. 2006 | USA | 5692 |  | Cross-sectional study | DSM IV | Alcohol abuse | 32.9 (3.0) | 3.1 | (2.3-4.1) |  |  |  |
|  |  |  |  |  |  | Alcohol dependence | 17.0(2.0) | 3.6 | (2.5-5.0) |  |  |  |
| **Study ID** | **Study Year** | **Sample Size** | **Sample Type** | **Study Design** | **Diagnostic Criteria** | **(Specific Comorbidity / Correlate)** | **pre% (standard error)** | **OR** | **95% CI** | **p-value** | **χ2** | **SE** |
| Al-Hamzawi et al.2012 Narrow | 2012 | 65 | Community | Cross-sectional survey | DSM IV | GAD | 39.2 (13.2) | 16 | 5.5-46.4 | <.0001 | 27.4 | 13.2 |
| Al-Hamzawi et al. 2012 Broad | 2012 | 77 | Community | Cross-sectional survey | DSM IV | GAD | 38.2 (11.7) | 15.7 | 6.1-40.5 | <0.0001 |  |  |
| Galbraith et al. (2018) | 2018 | 486 | Community | Cross-sectional survey | DSM 5 | GAD | 4.07（1.70） | 2.05 | 0.84-5.02 |  |  |  |
| Lee et al. (2016) | USA | IED: 42, PC: 50 | Clinical | Cross-sectional survey | DSM 5 | GAD | 0vs 4 | 0.12 | 0.006,2.33 | 0.061 |  |  |
| McLaughlin et al. (2012)- Broad | USA | 474 | Community | Cross-sectional survey | DSM IV | GAD | 2.6(1.0) | 0.7 | 0.3-1.7 |  |  |  |
| McLaughlin et al. (2012)- Narrow | USA | 343 | Community | Cross-sectional survey | DSM IV | GAD | 3.1(1.4) | 2.6 | 0.5-14.5 |  |  |  |
| Nickerson et al. (2012) | USA | 618 | Community | Cross-sectional survey | DSM IV | GAD |  | 2.61 | 1.91-3.58 |  |  |  |
| Ortega et al. (2008)-Lifetime | USA | 158 | Community | Cross-sectional survey | DSM IV | GAD |  | 2.58 | 1.41-4.72 |  |  |  |
| Ortega et al. (2008)-12 month | Mexico | 106 | Community | Cross-sectional survey | DSM IV | GAD |  | 3.29 | 3.29-7.98 |  |  |  |
| Pereira et al. (2021) | Brazil | 171 | Community | Cross-sectional survey | DSM 5 | GAD |  | 3.4 | 2.07-5.74 |  |  |  |
| Reardon et al. (2014) | USA | 56 | Clinical | Cross-sectional study | DSM 5 | GAD |  | 1.69 | 0.88-3.26 |  |  |  |
| Scott et al. (2016)-Lifetime | Cross-national (WMH) | 705 | Community | Cross-sectional study | DSM 5 | GAD | 20.8 (2.1) |  |  |  |  |  |
| Scott et al. (2016)- 12 months | Cross-national (WMH) | 357 | Community | Cross-sectional study | DSM 5 | GAD | 19.1 (2.8) |  |  |  |  |  |
| Scott et al. 2020 | Cross-national (WMH) | 651 | Community | Cross-sectional study | DSM 5 | GAD | 20.9(2.1) | 6.6 | 5.0-8.7 |  |  |  |
| Tay et al. (2022) | Malaysia | 279 | Community | Cross-sectional study | DSM 5 | GAD |  |  |  |  |  |  |
| Yoshimasu & Kawakami (2011)-Male | Japan | 52 | Community | Cross-sectional study | DSM IV | GAD | 5.90% | 4.9 | 1.4-17.1 |  |  |  |
| Yoshimasu & Kawakami (2011)-female | Japan | 28 | Community | Cross-sectional study | DSM IV | GAD | 30.3 | 7.2 | 2.4-21.9 |  |  |  |
| Kesssler et al. 2006 | USA | 5692 | Community | Cross-sectional study | DSM IV | GAD | 18.7 (1.8) | 3.6 | (2.8-4.7) |  |  |  |
| **Study ID** | **Study Year** | **Sample Size** | **Sample Type** | **Study Design** | **Diagnostic Criteria** | **(Specific Comorbidity / Correlate)** | **pre% (standard error)** | **OR** | **95% CI** | **p-value** |  |  |
| Al-Hamzawi et al.2012 Narrow | 2012 | 65 | Community | Cross-sectional survey | DSM IV | MDD | 21.3 （7.2） | 4 | 1.8-9.1 | 0.001 |  |  |
| Al-Hamzawi et al. 2012 Broad | 2012 | 77 | Community | Cross-sectional survey | DSM IV | MDD | 21.9 (6.7) | 4.2 | 2.0-8.7 | 0 |  |  |
| Coccaro. 2019-NCS-AS | 2019 | 486 | Community | Cross-sectional survey | DSM 5 | Depressive Disorder | 19.0%(IED) vs 7.1% | 2.14 | 1.64-2.80 | <0.001 |  |  |
| Coccaro. 2019-NCS-R | 2019 | 463 | Community | Cross-sectional survey | DSM 5 | Depressive Disorder | 22.3% vs 7.5% | 1.55 | 1.07-2.55 | <0.05 |  |  |
| Coccaro. 2019-Clinical | 2019 | 744 | Clinical | Cross-sectional survey | DSM 5 | Depressive Disorder | 21.1 % vs 10.5% | 1.63 | 1.20-2.20 |  |  |  |
| Gelegen & Tamam (2018) | 2018 | 68 | Clinical | Cross-sectional survey | DSM 5 | Depression | 32.4 | 0.92 | 0.53-1.60 |  |  |  |
| Lee et al. (2016) | USA | IED: 42, PC: 50 | Clinical | Cross-sectional survey | DSM 5 | MDD | 7 vs 0 | 21.34 | 1.18, 385.0 | 0.003 |  |  |
| McLaughlin et al. (2012)- Broad | USA | 474 | Community | Cross-sectional survey | DSM IV | MDD | 21.6 (2.2) | 1.1 | 0.8-1.5 |  |  |  |
| McLaughlin et al. (2012)- Narrow | USA | 343 | Community | Cross-sectional survey | DSM IV | MDD | 23.0(3.1) | 1.5 | 0.7-3.2 |  |  |  |
| Ortega et al. (2008)-Lifetime | USA | 158 | Community | Cross-sectional survey | DSM IV | MDD |  | 3.21 | 1.94-5.20 |  |  |  |
| Ortega et al. (2008)-12 month | Mexico | 106 | Community | Cross-sectional survey | DSM IV | MDD |  | 4.22 | 2.34-7.63 |  |  |  |
| Pereira et al. (2021) | Brazil | 171 | Community | Cross-sectional survey | DSM 5 | MDD |  | 2.8 | 2.21-3.48 |  |  |  |
| Puhalla et al. (2020) | USA | 533 | Clinical | Cross-sectional survey | DSM 5 | Depression |  | 0.54 | 0.38-0.77 |  |  |  |
| Reardon et al. (2014) | USA | 56 | Clinical | Cross-sectional study | DSM 5 | MDD |  | 1.73 | 0.87-3.45 |  |  |  |
| Scott et al. (2016)-Lifetime | Cross-national (WMH) | 705 | Community | Cross-sectional study | DSM 5 | MDD | 35.2 (2.5) |  |  |  |  |  |
| Scott et al. (2016)- 12 months | Cross-national (WMH) | 357 | Community | Cross-sectional study | DSM 5 | MDD | 25.2 (2.9) |  |  |  |  |  |
| Scott et al. 2020 | Cross-national (WMH) | 651 | Community | Cross-sectional study | DSM 5 | MDD | 26.4(2.4) | 2.7 | 2.1-3.5 |  |  |  |
| Shevidi et al. (2023) | USA | 393 | Clinical | Cross-sectional study | DSM 5 | Depression | 68% | 1.5 | 1.08-2.09 |  |  |  |
| Tay et al. (2022) | Malaysia | 279 | Community | Cross-sectional study | DSM 5 | MDD | 67.60% | 3.1 | 1.87-5.14 | p=0.002 |  |  |
| Tay et al. (2015) | Papua New Guinea | 50 | Community | Cross-sectional study | DSM 5 | MDD |  | 3.1 | 1.87-5.14 |  |  |  |
| Yoshimasu & Kawakami (2011)-Male | Japan | 52 | Community | Cross-sectional study | DSM IV | MDD | 15.20% | 4.8 | 2.1-10.7 |  |  |  |
| Yoshimasu & Kawakami (2011)-female | Japan | 28 | Community | Cross-sectional study | DSM IV | MDD | 13.7 | 1.6 | 0.5-5.0 |  |  |  |
| Kesssler et al. 2006 | USA | 5692 |  | Cross-sectional study | DSM IV | MDD | 37.3 (SE) | 2.8 | (2.2-3.6) |  |  |  |
| **Study ID** | **Study Year** | **Sample Size** | **Sample Type** | **Study Design** | **Diagnostic Criteria** | **(Specific Comorbidity / Correlate)** | **pre% (standard error)** | **OR** | **95% CI** | **p-value** | **χ2** | **SE** |
| Al-Hamzawi et al.2012 Narrow | 2012 | 65 | Community | Cross-sectional survey | DSM IV | Panic Disorder | 2.6 (0.8) | 2 | 0.9-4.4 | 0.07 | 3.2 | 0.8 |
| Al-Hamzawi et al. 2012 Broad | 2012 | 77 | Community | Cross-sectional survey | DSM IV | Panic Disorder | 2.5 (0.7) | 1.9 | 0.9-4.1 | 0.09 |  |  |
| Galbraith et al. (2018) | 2018 | 486 | Community | Cross-sectional survey | DSM 5 | Panic Disorder | 6.84 （1.74） | 4.13 | 2.30-7.39 | p<.05 |  |  |
| Lee et al. (2016) | USA | IED: 42, PC: 50 | Clinical | Cross-sectional survey | DSM 5 | Panic Disorder | 0 vs 2 | 0.23 | 0.01, 4.87 | 0.19 |  |  |
| McLaughlin et al. (2012)- Broad | USA | 474 | Community | Cross-sectional survey | DSM IV | Panic Disorder | 7.2(2.2) | 3.8 | 1.8-7.9 |  |  |  |
| McLaughlin et al. (2012)- Narrow | USA | 343 | Community | Cross-sectional survey | DSM IV | Panic Disorder | 8.6 (2.9) | 2.5 | 0.3-24.0 |  |  |  |
| Ortega et al. (2008)-Lifetime | USA | 158 | Community | Cross-sectional survey | DSM IV | Panic Disorder |  | 6.1 | 2.49-14.92 |  |  |  |
| Ortega et al. (2008)-12 month | Mexico | 106 | Community | Cross-sectional survey | DSM IV | Panic Disorder |  | 10.35 | 3.07-34.87 |  |  |  |
| Pereira et al. (2021) | Brazil | 171 | Community | Cross-sectional survey | DSM 5 | Panic Disorder |  | 4.7 | 2.43-9.17 |  |  |  |
| Reardon et al. (2014) | USA | 56 | Clinical | Cross-sectional study | DSM 5 | Panic Disorder |  | 1.32 | 0.67-2.59 |  |  |  |
| Scott et al. (2016)-Lifetime | Cross-national (WMH) | 705 | Community | Cross-sectional study | DSM 5 | Panic Disorder | 9.5 (1.2) |  |  |  |  |  |
| Scott et al. (2016)- 12 months | Cross-national (WMH) | 357 | Community | Cross-sectional study | DSM 5 | Panic Disorder | 8.3 (1.5) |  |  |  |  |  |
| Scott et al. 2020 | Cross-national (WMH) | 651 | Community | Cross-sectional study | DSM 5 | Panic Disorder | 9.6(1.2) | 4.6 | 3.4-6.3 |  |  |  |
| Kesssler et al. 2006 | USA | 5692 |  | Cross-sectional study | DSM IV | Panic Disorder | 11.9 (1.6) | 3.3 | (2.2-4.8) |  |  |  |
| **Study ID** | **Study Year** | **Sample Size (with IED)** | **Study Design** | **Diagnostic Criteria** | **(Specific Comorbidity / Correlate)** | **pre% (standard error)** | **OR** | **95% CI** | **p-value** | **Sample type** |  |  |
| Al-Hamzawi et al.2012 Narrow | 2012 | 65 | Cross-sectional survey | DSM IV | PTSD |  | 5.8 | 2.3-14.5 | 0 | community |  |  |
| Al-Hamzawi et al. 2012 Broad | 2012 | 77 | Cross-sectional survey | DSM IV | PTSD |  | 6.9 | 2.9-16.1 | <0.0001 | community |  |  |
| Coccaro. 2019-NCS-AS | 2019 | 486 | Cross-sectional survey | DSM 5 | PTSD |  | 1.56 | 1.06-2.31 | <0.001 | community |  |  |
| Coccaro. 2019-NCS-R | 2019 | 463 | Cross-sectional survey | DSM 5 | PTSD |  | 2.04 | 1.29-3.24 | <0.05 | community |  |  |
| Coccaro. 2019-Clinical | 2019 | 744 | Cross-sectional survey | DSM 5 | PTSD |  | 3.42 | 2.09-5.59 |  | clinical |  |  |
| Lee et al. (2016) | USA | 42 | Cross-sectional survey | DSM 5 | PTSD | 3 vs 1 | 3.77 | 0.38,37.66 | 0.228 | clinical |  |  |
| McLaughlin et al. (2012)- Broad | USA | 474 | Cross-sectional survey | DSM IV | PTSD |  | 1.4 | 0.7-2.8 |  | community |  |  |
| McLaughlin et al. (2012)- Narrow | USA | 343 | Cross-sectional survey | DSM IV | PTSD |  | 0.7 | 0.1-7.1 |  | community |  |  |
| Nickerson et al. (2012) | USA | 618 | Cross-sectional survey | DSM IV | PTSD |  | 1.67 | 1.23-2.26 |  | community |  |  |
| Ortega et al. (2008)-Lifetime | Mexico | 158 | Cross-sectional survey | DSM IV | PTSD |  | 4.46 | 2.43-8.18 |  | community |  |  |
| Ortega et al. (2008)-12 month | Mexico | 106 | Cross-sectional survey | DSM IV | PTSD |  | 11.55 | 5.98-22.30 |  | community |  |  |
| Pereira et al. (2021) | Brazil | 171 | Cross-sectional survey | DSM 5 | PTSD |  | 2.8 | 1.32-6.07 |  | community |  |  |
| Puhalla et al. (2020) | USA | 533 | Cross-sectional survey | DSM 5 | PTSD |  | 0.64 | 0.36-1.12 |  | clinical |  |  |
| Reardon et al. (2014) | USA | 56 | Cross-sectional survey | DSM 5 | PTSD |  | 2.63 | 1.29-5.35 |  | clinical |  |  |
| Scott et al. (2016)-Lifetime | Cross-national (WMH) | 705 | Cross-sectional survey | DSM 5 | PTSD | 15.5 (1.5) |  |  |  | community |  |  |
| Scott et al. (2016)- 12 months | Cross-national (WMH) | 357 | Cross-sectional survey | DSM 5 | PTSD | 11.0 (1.7) |  |  |  | community |  |  |
| Scott et al. 2020 | Cross-national (WMH) | 651 | Cross-sectional survey | DSM 5 | PTSD | 16.4(1.7) | 4.9 | 3.8-6.3 |  | community |  |  |
| Silove et al. 2015 | Timor-Leste | 186 | Cross-sectional survey | DSM 5 | PTSD |  | 2.3 | 0-0.90 |  | community |  |  |
| Tay et al. (2022) | Malaysia | 279 | Cross-sectional survey | DSM 5 | PTSD | 42.30% | 2.73 | 1.69-4.42 | p=0.002 | community |  |  |
| Tay et al. (2015) | Papua New Guinea | 50 | Cross-sectional survey | DSM 5 | PTSD |  | 2.73 | 1.69-4.42 |  | community |  |  |
| Kesssler et al. 2006 | USA | 5692 |  | DSM IV | PTSD | 15.2 (1.5) | 3 | (2.3-4.1) |  | community |  |  |
| **Study ID** | **Study Year** | **Sample Size** | **Sample Type** | **Study Design** | **Diagnostic Criteria** | **(Specific Comorbidity / Correlate)** | **pre% (standard error)** | **OR** | **95% CI** |  |  |  |
| Al-Hamzawi et al.2012 Narrow | 2012 | 65 | Community | Cross-sectional survey | DSM IV | Social phobia | 5.2 (3.5) | 7.9 | 1.5-41.1 |  |  |  |
| Al-Hamzawi et al. 2012 Broad | 2012 | 77 | Community | Cross-sectional survey | DSM IV | Social phobia | 4.6 (3.1) | 7 | 1.4-35.5 |  |  |  |
| Galbraith et al. (2018) | 2018 | 486 | Community | Cross-sectional survey | DSM 5 | Social phobia | 14.87（2.04） | 2.18 | 1.57-3.03 |  |  |  |
| McLaughlin et al. (2012)- Broad | USA | 474 | Community | Cross-sectional survey | DSM IV | Social phobia | 16.0(2.9) | 2.2 | 1.3-3.7 |  |  |  |
| McLaughlin et al. (2012)- Narrow | USA | 343 | Community | Cross-sectional survey | DSM IV | Social phobia | 16.6(4.1) | 1.3 | 0.5-3.3 |  |  |  |
| Ortega et al. (2008)-Lifetime | USA | 158 | Community | Cross-sectional survey | DSM IV | Social phobia | | 3.31 | 1.87-5.87 |  |  |  |
| Ortega et al. (2008)-12 month | Mexico | 106 | Community | Cross-sectional survey | DSM IV | Social phobia | | 4.39 | 2.17-8.88 |  |  |  |
| Pereira et al. (2021) | Brazil | 171 | Community | Cross-sectional survey | DSM 5 | Social phobia | | 3.5 | 2.27-5.50 |  |  |  |
| Reardon et al. (2014) | USA | 56 | Clinical | Cross-sectional study | DSM 5 | Social phobia | | 1.13 | 0.57-2.24 |  |  |  |
| Scott et al. (2016)-Lifetime | Cross-national (WMH) | 705 | Community | Cross-sectional study | DSM 5 | Social phobia | 23.1 (1.9) |  |  |  |  |  |
| Scott et al. (2016)- 12 months | Cross-national (WMH) | 357 | Community | Cross-sectional study | DSM 5 | Social phobia | 18.8 (2.5) |  |  |  |  |  |
| Scott et al. 2020 | Cross-national (WMH) | 651 | Community | Cross-sectional study | DSM 5 | Social phobia | 23.4(2.0) | 4.9 | 3.9-6.3 |  |  |  |
| Kesssler et al. 2006 | USA | 5692 |  | Cross-sectional study | DSM IV | Social phobia | 28.3 (1.5) | 3.1 | (2.5-3.7) |  |  |  |
